# IGenomic answers for children: Dynamic analyses of >1000 pediatric rare disease genomes

**DOI:** 10.1101/2021.10.07.21264628

**Authors:** Ana SA Cohen, Emily G Farrow, Ahmed T Abdelmoity, Joseph T Alaimo, Shivarajan M Amudhavalli, John T Anderson, Lalit Bansal, Lauren Bartik, Primo Baybayan, Bradley Belden, Courtney D Berrios, Rebecca L Biswell, Pawel Buczkowicz, Orion Buske, Shreyasee Chakraborty, Warren A Cheung, Keith A Coffman, Ashley M Cooper, Laura A Cross, Thomas Curran, Thuy Tien T Dang, Mary M Elfrink, Kendra L Engleman, Erin D Fecske, Cynthia Fieser, Keely Fitzgerald, Emily A Fleming, Randi N Gadea, Jennifer L Gannon, Rose N Gelineau-Morel, Margaret Gibson, Jeffrey Goldstein, Elin Grundberg, Kelsee Halpin, Brian S Harvey, Bryce A Heese, Wendy Hein, Suzanne M Herd, Susan S Hughes, Mohammed Ilyas, Jill Jacobson, Janda L Jenkins, Shao Jiang, Jeffrey J Johnston, Kathryn Keeler, Jonas Korlach, Jennifer Kussmann, Christine Lambert, Caitlin Lawson, Jean-Baptiste Le Pichon, Steve Leeder, Vicki C Little, Daniel A Louiselle, Michael Lypka, Brittany D McDonald, Neil Miller, Ann Modrcin, Annapoorna Nair, Shelby H Neal, Christopher M Oermann, Donna M Pacicca, Kailash Pawar, Nyshele L Posey, Nigel Price, Laura MB Puckett, Julio F Quezada, Nikita Raje, William J Rowell, Eric T Rush, Venkatesh Sampath, Carol J Saunders, Caitlin Schwager, Richard M Schwend, Elizabeth Shaffer, Craig Smail, Sarah Soden, Meghan E Strenk, Bonnie R Sullivan, Brooke R Sweeney, Jade B Tam-Williams, Adam M Walter, Holly Welsh, Aaron M Wenger, Laurel K Willig, Yun Yan, Scott T Younger, Dihong Zhou, Tricia N Zion, Isabelle Thiffault, Tomi Pastinen

**Affiliations:** Genomic Medicine Center, Department of Pathology and Laboratory Medicine, Children’s Mercy Kansas City; University of Missouri-Kansas City School of Medicine; Department of Genetics, Children’s Mercy Kansas City; Department of Pediatrics, Children’s Mercy Kansas City; PhenoTips; Pacific Biosciences

## Abstract

**PURPOSE:** To provide comprehensive diagnostic and candidate analyses in a pediatric rare disease cohort through the Genomic Answers for Kids (GA4K) program.

**METHODS:** Extensive analyses of 960 families with suspected genetic disorders including short-read exome (ES) and genome sequencing (srGS); PacBio HiFi long-read GS (HiFi-GS); variant calling for small-nucleotide (SNV), structural (SV) and repeat variants; and machine-learning variant prioritization. Structured phenotypes, prioritized variants and pedigrees are stored in PhenoTips database, with data sharing through controlled access (dbGAP).

**RESULTS:** Diagnostic rates ranged from 11% for cases with prior negative genetic tests to 34.5% in naïve patients. Incorporating SVs from GS added up to 13% of new diagnoses in previously unsolved cases. HiFi-GS yielded increased discovery rate with >4-fold more rare coding SVs than srGS. Variants and genes of unknown significance (VUS/GUS) remain the most common finding (58% of non-diagnostic cases).

**CONCLUSION:** Computational prioritization is efficient for diagnostic SNVs. Thorough identification of non-SNVs remains challenging and is partly mitigated by HiFi-GS sequencing. Importantly, community research is supported by sharing real-time data to accelerate gene validation, and by providing HiFi variant (SNV/SV) resources from >1,000 human alleles to facilitate implementation of new sequencing platforms for rare disease diagnoses.

## INTRODUCTION

The Children’s Mercy Research Institute (CMRI) in Kansas City established a large-scale genomic disease program named “Genomic Answers for Kids” (GA4K) to expand diagnostic capabilities and catalog rare disease genomes and phenotypes within a healthcare system. Broad recruitment across all pediatric rare diseases resulted in most patients entering the study either with negative or no prior genetic testing. Recent studies have shown >10% rate of new findings upon reanalysis of exome or genome sequencing data in patients with a history of negative genetic testing.^1-4^ The predominant factors in identifying new diagnoses were recent publications establishing novel gene-disease associations, often through data-sharing efforts such as GeneMatcher (upgrade from ‘gene of uncertain significance’ or GUS), or expanding the phenotypic spectrum of established disease genes (upgrade from ‘variant of uncertain significance’ or VUS).^1,3,5^ The next most helpful strategy to increase diagnostic yield was the incorporation of sequencing data from additional family members, particularly for singletons.^4^ Further, given the continued advances in technology and expanded availability of public data, samples sequenced and/or analyzed >3-5 years prior may also benefit from re-sequencing to enhance coverage and/or re-pipelining to incorporate improved filtering methods and more extensive population data.^3,6^

The variable success in analyses/re-analyses is largely explained by patient ascertainment and testing schemes, though differing variant prioritization strategies are also likely to play a role. Specifically, depending on the relative weight placed on inheritance, variant-effect properties, and the identity/function of the gene harboring the rare variant, the ranking of candidate variants may yield very different results. Multiple machine-learning tools have emerged to balance the variant/locus characteristics in an attempt to systematically extract optimal candidate prioritization.^7^ The integration of such tools in rare disease molecular analyses has been demonstrated by several centers primarily for small, selected cohorts.^8-12^ The universal feature is the patient’s phenotype coded through human phenotype ontology (HPO) terms as a basis for prioritization, followed by the deployment of variable ranking algorithms.^13^ However, the utility of incorporating such tools for a systematic first-pass analysis of patient data within a large, unselected, and phenotypically diverse pediatric rare disease diagnostic setting is unknown.

While variant prioritization strategies continue to improve, the choice of technology in genome-wide sequencing and primary data processing strategy have remained comparatively stable, despite missing some variant types including structural variation.^14-16^ At our center, short-read genome sequencing (srGS) performed similarly to exome sequencing (ES) in the diagnostic evaluation of suspected pediatric genetic disease on the same Illumina platform.^17^ However, alternative platforms have the potential to reduce uncertainty of chemistry-dependent errors and omissions, and scalable alternatives have emerged for short-read PCR-free genomes such as DNA NanoBall (DNB) sequencing.^18^ Further, long-read GS (lrGS) has been shown to detect variants missed by short-read sequencing, specifically complex structural variants including inversions and inverted duplications, as well as repeat expansions and variants in difficult-to-map regions.^19^ In addition, lrGS also has the potential to resolve phasing of variants in autosomal recessive genes when parental samples are unavailable. Recent technological advances in long-read platforms enable the consideration of lrGS for unsolved rare diseases.^20^

Herein we leveraged a large scale pediatric genomic medicine program with real-time return of results to explore automation of variant prioritization and expert clinical interpretation, as well as the re-testing of prior negative exomes at a scale that has not been previously reported. The results from the analyses of over 1000 rare disease patients highlight the utility of systematic variant prioritization, identify variants in ‘blind spots” associated with current technologies, and underscore the imperative for improved sharing strategies of suggestive results across rare disease programs and cohorts.^21^

## MATERIALS and METHODS

Detailed methods are described in the Supplementary text online. All analyses were completed on GRCh38.

### Cohort

The case cohort described includes 1083 affected patients from 960 families, with a total of 2,957 sequenced individuals collectively (detailed in Supplementary Tables S1 and S2). Cases included 595 males and 488 females, ages 1 to 55 years old (older individuals were typically ascertained as follow-up from affected child). Of these, 158 (14.6%) were singletons whereas the remaining 955 had at least one additional family member sequenced. Patients were referred from 22 different specialties, with the largest proportion nominated by Clinical Genetics (47.7%), followed by Neurology (22.9%). Given the broad referral pool, we acknowledge limitations in the ethnic diversity of this population that may reflect systemic healthcare issues; these will be addressed directly in future studies. A continuum of pediatric conditions is represented, ranging from congenital anomalies to more subtle neurological and neurobehavioral clinical presentations later in childhood. Of the 1083 patients, 125 entered the study with a known genetic diagnosis, as the program is building an inclusive rare disease genome resource with solved cases serving to benchmark new methods and processes. Phenotypes were manually extracted from the medical records and primary features recorded in PhenoTips utilizing HPO terminology.^13,22^ These structured data were used for automated prioritization tools, whereas expert review used the complete clinical notes for variant prioritization and interpretation.

### Short-read exome and genome sequencing (ES/srGS)

Exome libraries were prepared according to manufacturer’s standard protocols using the Illumina TruSeq PCR-Free library preparation kit (Illumina, San Diego, CA) with 10 cycles of PCR, followed by enrichment with the IDT xGen Exome Research Panel v2, with additional spike-in oligos (Integrated DNA Technologies, Coralville, IA) to capture the mitochondrial genome and dispersed genomic regions for CNV detection.^23^ PCR-Free genome libraries were prepared according to manufacturer’s standard protocols for Illumina TruSeq library preparation.

### MGI sequencing (srGS)

Genome sequencing libraries were constructed using the MGIEasy Universal DNA Library Prep Set (MGI, Shenzhen, Guangdong, China) according to manufacturer’s standard protocols. srGS was performed on an MGI DNBSEQ-G400.

### PacBio HiFi long-read sequencing (HiFi-GS) and analysis

DNA was sheared to a target size of 14 kb using the Diagenode Megaruptor3 (Diagenode, Liege, Belgium). SMRTbell libraries were prepared with the SMRTbell Express Template Prep Kit 2.0 (100-938-900, Pacific Biosciences, Menlo Park, CA) following the manufacturer’s standard protocol (101-693-800) with modifications described in the Supplementary methods. Libraries were sequenced on the Sequel IIe Systems using the Sequel II Binding Kit 2.0 (101-842-900) or 2.2 (102-089-000) and Sequel II Sequencing Kit 2.0 (101-820-200) with 30 hr movies/SMRT cell. 175 samples were sequenced to a target of >25X coverage; 297 samples were sequenced on 1 SMRT Cell (average: 10X coverage).

Read mapping, variant calling, and genome assembly were performed using a Snakemake workflow. HiFi reads were mapped with pbmm2 v1.4.0 and structural variants were called with pbsv 2.4.0 Small variants were called with DeepVariant 1.0 following DeepVariant best practices for PacBio reads. ^23^ *De novo* assembly was performed with hifiasm v0.9-r289 using default parameters.^24^

Structural variant call sets were compared using svpack match which considers two SV calls to match when the variants are of the same type (considering INS and DUP to be the same), nearby (start position difference ≤ 100 bp), and similar size (size difference ≤ 100 bp). To systematically evaluate expansions at known pathogenic tandem repeat loci, tandem-genotypes was used to count the length of tandem repeats in HiFi reads for each sample.^25^ As long [GA]-rich repeats have been noted to have lower coverage in HiFi reads, a complementary system was setup to identify haplotypes with coverage dropouts at the known pathogenic tandem repeat loci.^26^ At each locus, the number of reads that span the repeat region were counted per haplotype (based on a Whatshap-haplotagged BAM from phased SNVs).^27^ A coverage dropout was identified as a locus with fewer than 2 spanning reads in a haplotype.

Joint calling of structural and small variants was also completed for HiFi-GS. A multi-sample structural variant callset was produced by merging single-sample pbsv callsets with JASMINE v1.1.4.^28^ A multi-sample small variant callset was produced by running GLnexus v1.2.7 on all single-sample DeepVariant gVCF files using glnexus_cli --config DeepVariant_unfiltered and converting the resulting BCF to VCF with bcftools view v1.10.^29^

### Analyses and variant prioritization pipeline

Figure 1 depicts an overview of the sequence processing, variant calling and interpretation pipeline. Re-analysis was carried out using ES/srGS data in parallel. Exomiser v12.1 (data version 2102) and AMELIE v3.1.0 were applied for variant prioritization and highly ranked variants were manually reviewed and flagged for expert interpretation.^30,31^ An additional sequencing platform using srGS was tested in a subset of trios (MGI), whereas long-read HiFi-GS (PacBio) was predominantly deployed for cases without diagnosis after srGS. Finally, an early phase of the study employed 10X-linked read GS, predominantly in singleton cases (see Supplementary methods). Supplementary Tables S3 summarizes the different types of data generated for the cohort.

**Figure 1.**
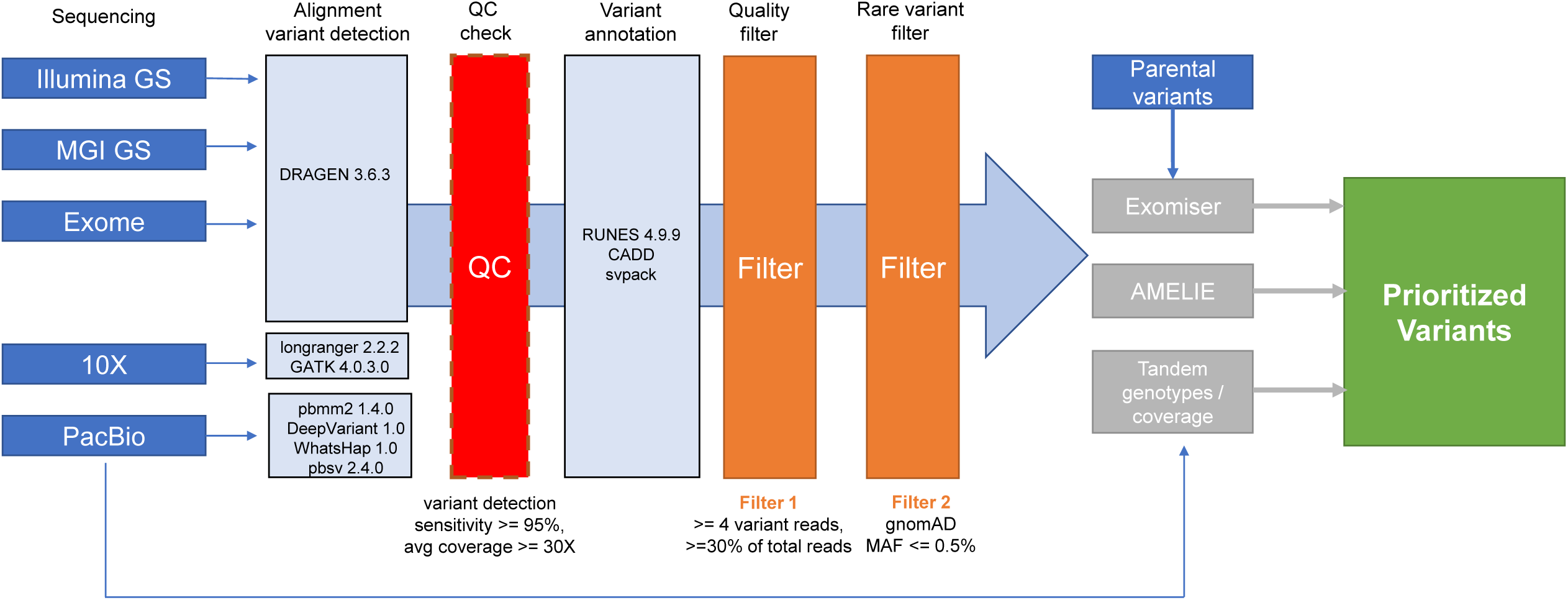
Genomic Answers for Kids (GA4K) pipeline. Overview of sequencing, variant calling and variant prioritization pipeline. Sequencing included exome sequencing as well as genome sequencing through multiple technologies (Illumina, MGI, and 10X for short-reads, and PacBio for long-reads). Standard quality control (QC) and filtering were applied. Variant prioritization relied on inheritance pattern and AI tools (Exomiser/Amelie) as well as tandem genotypes.

Annotation of structural variants (SVs) for disease relevance utilized both frequency (MAF <1%) in a sequence modality specific, local, SV warehouse, and focused on overlap with OMIM morbid genes, followed by manual curation to interpret the validity of candidate SV calls, as well as relevance in context of the phenotype/known transmission of disease at locus.

### Clinical validation of research results

Variants identified through research sequencing were reviewed according to ACMG criteria; pathogenic and likely pathogenic variants related to the disease phenotype were confirmed in the Children’s Mercy CLIA-certified laboratory through best applicable validated methods and reported clinically in real-time for incorporation into clinical management.^32^

## RESULTS

### Machine-assisted interpretation

A combination of two publicly available tools was implemented to aid with variant prioritization: Exomiser and AMELIE.^30,31^ Both tools (E/A) rely on structured phenotyping (with HPO terms) but apply algorithms that explore different features of variants/genes (see Supplementary methods). Therefore, we hypothesized that the combination would improve speed and accuracy of analysis. To test the efficacy of these tools, we first reviewed the combined top 50 ranked E/A candidate variant list for cases with known molecular diagnoses at study entry (n=125), with knowledge of the phenotype of each proband but blinded to the original genetic results. Of these, 88 had diagnostic SNVs serving as a positive control set (other known diagnoses included aneuploidies, microdeletions/duplications, repeat expansions, or special cases such as *SMN1*/2 variants not in the scope of exome or genome interpretation provided here, and described in Figure 2a as “other mechanism”). The causative variant was ranked by E/A in 84 (95.5%) of “positive control” cases. Three of the four cases for which the diagnostic variant was not ranked had deep intronic pathogenic variants, a recognized limitation of E/A prioritization; therefore, only one diagnostic coding variant was missed in this subset of cases.

**Figure 2.**
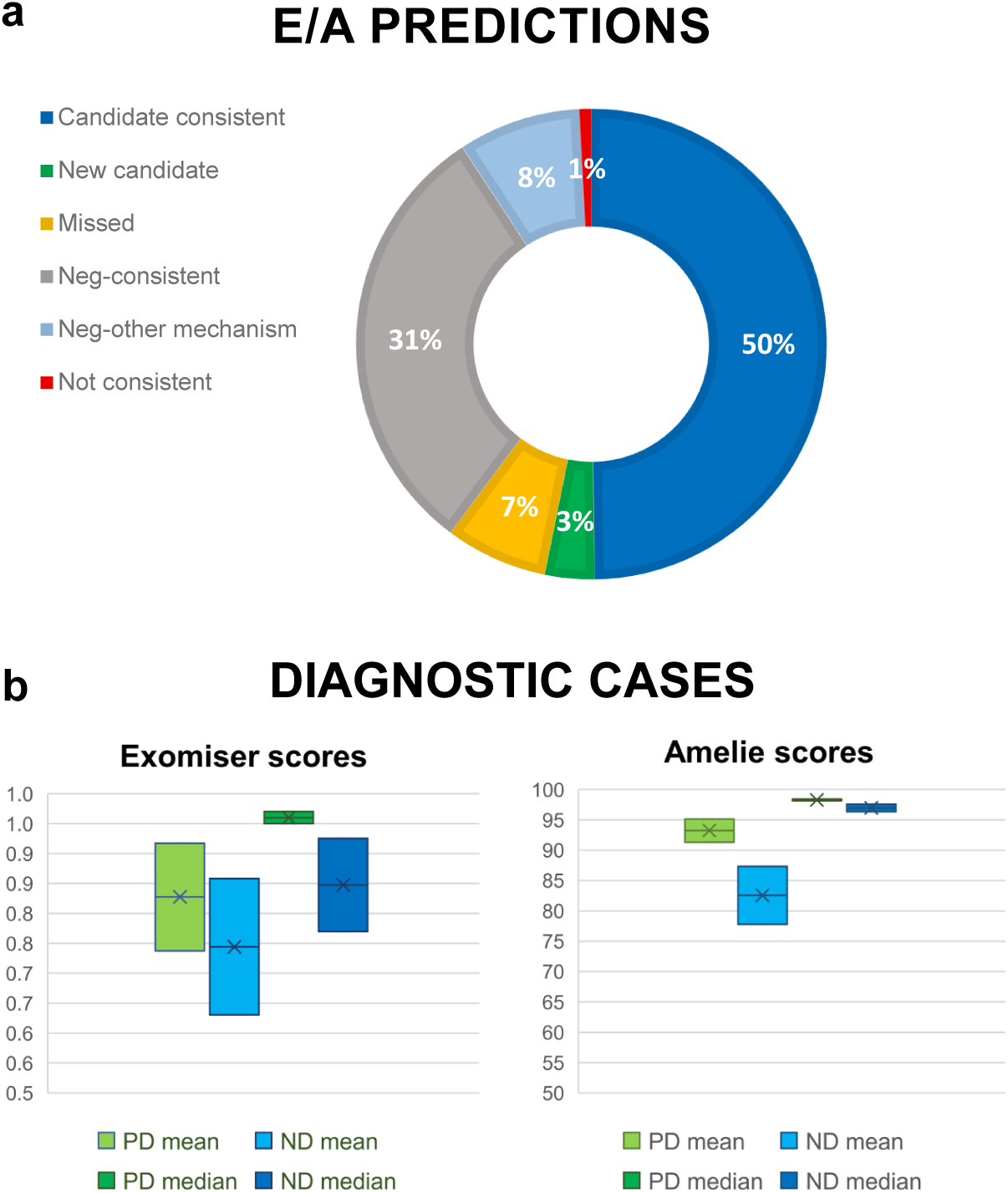
Variant prioritization tools showed great concordance with expert analysis. (**a**) Distribution of Exomiser/Amelie (E/A) predictions for all 1083 patients. Prioritization was deemed concordant when the main candidate variant was concordant with expert review (“candidate consistent”), negative by both E/A and expert review (“Neg-consistent”), or when the causative variant was not an SNV and therefore not expected to be ranked by E/A (“Neg-other mechanism), totaling almost 89%. Prioritization was deemed non concordant when a different candidate variant was highly ranked (“Not consistent”) or when the top candidate was not ranked/very low ranked (“Missed”), totaling about 8%. Finally, approximately 3% had a new strong candidate variant prioritized by E/A that was missed by expert review. (**b**) The distribution of E/A scores in shown for cases with known diagnosis at enrollment (“PD”) and new diagnosis (“ND”). Exomiser scores range from 0 to 1, with 1 being the highest/best match. Amelie scores range from 0 to 100, with 100 being the highest/best match. Median is shown to illustrate the shift in mean due to a minority of missed rankings (when diagnostic variant was not ranked the lowest score was used).

Expanding the strategy to the entire dataset and comparing with expert review where variants were prioritized based on multiple criteria (zygosity, segregation, population frequency, gene function, etc.) as we previously described, variant prioritization was concordant in ∼49.8% of 1083 cases (Figure 2a), meaning that the top variants selected from the combined E/A files were consistent with those identified by expert review (score distribution is illustrated in Figure 2b).^33^ No strong E/A candidates were identified in ∼8.4% of cases which were positive for a variant that would not have been annotated by these tools (such as copy number variants, deep intronic variants, structural variants, repeat expansions, etc). Moreover, ∼30.6% of cases were deemed negative by both expert analysis and combined E/A ranking, giving us an overall consistency of ∼88.7% (see Figure 2a). Importantly, in ∼3.4% of cases (n=37), these tools pointed us towards new candidates that may not have otherwise been considered.

### Diagnostic yields stratified by earlier testing history

Of the 958/1083 patients (88.5%) that entered the study without a prior diagnosis, the largest group consisted of patients with an earlier negative genetic testing history (584/958), either by ES, srGS or panel testing. New ES and sr/lrGS with (re)analysis yielded definitive diagnoses for 64/584 cases (11%). A smaller group of patients, referred to the research study and to clinical ES in parallel, achieved a diagnostic rate of 71/206 cases (34.5%), and among patients that had no clinical genetic testing approved/ordered, the diagnostic rate was 34/168 (20.2%). Various modes of re-interpretation success are exemplified in Table 1 (and illustrated in Supplementary Figures S1-S8). We note that 8/64 of previously tested but negative cases were diagnosed by analyses of GS when ES analyses were negative, suggesting that among cohorts of ES tested patients the contribution of GS can be >10% of achievable diagnoses. Most stem from SVs with intronic breakpoints. Among previously untested patients, GS was required to solve pathogenic variation not detected by ES in 5/90 of diagnostic cases (6%) due to intronic variation, small deletions difficult to detect with ES, repeats expansion disorders, and disease associated non-coding RNAs not covered in the exome capture.

**Table 1.**
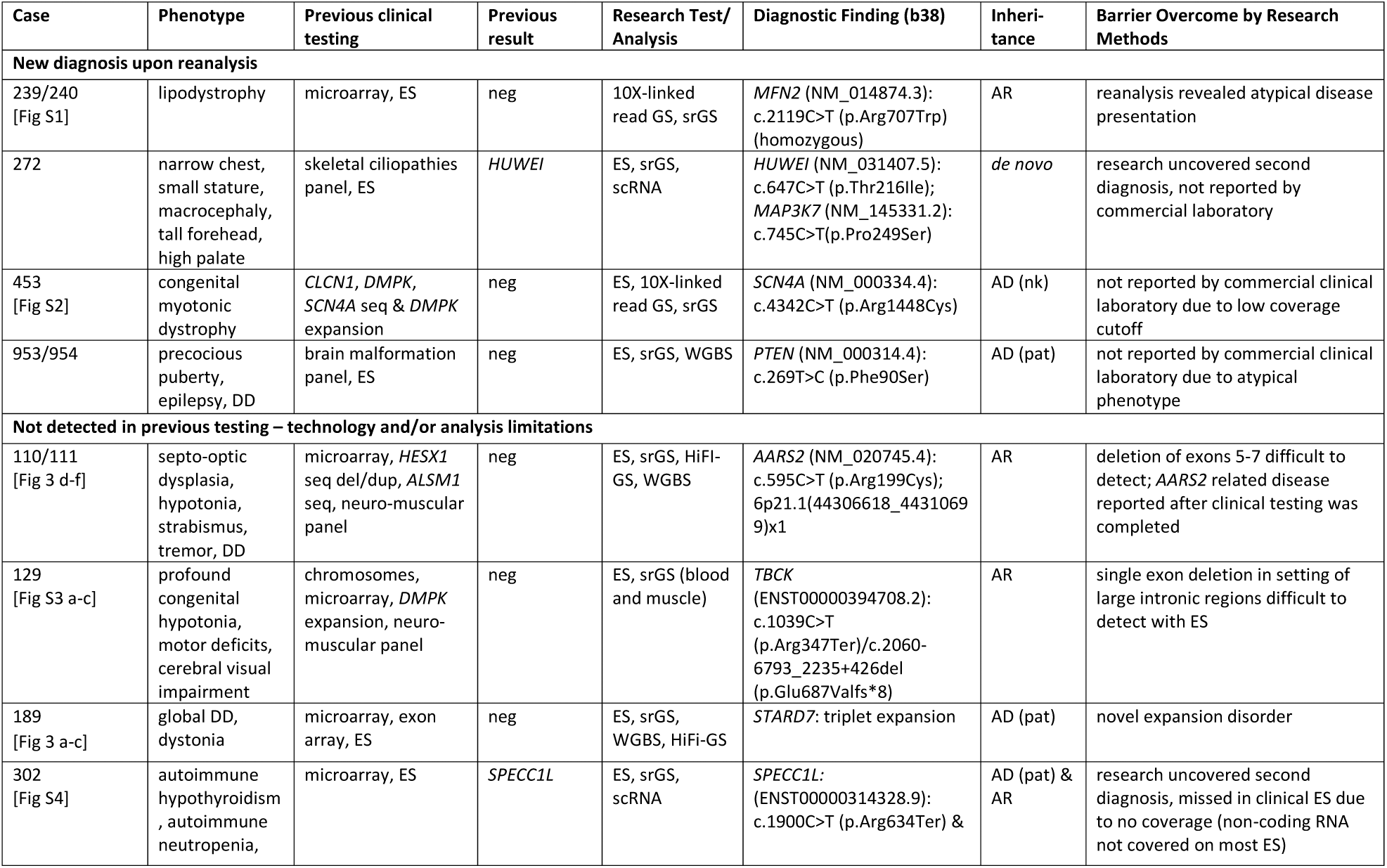
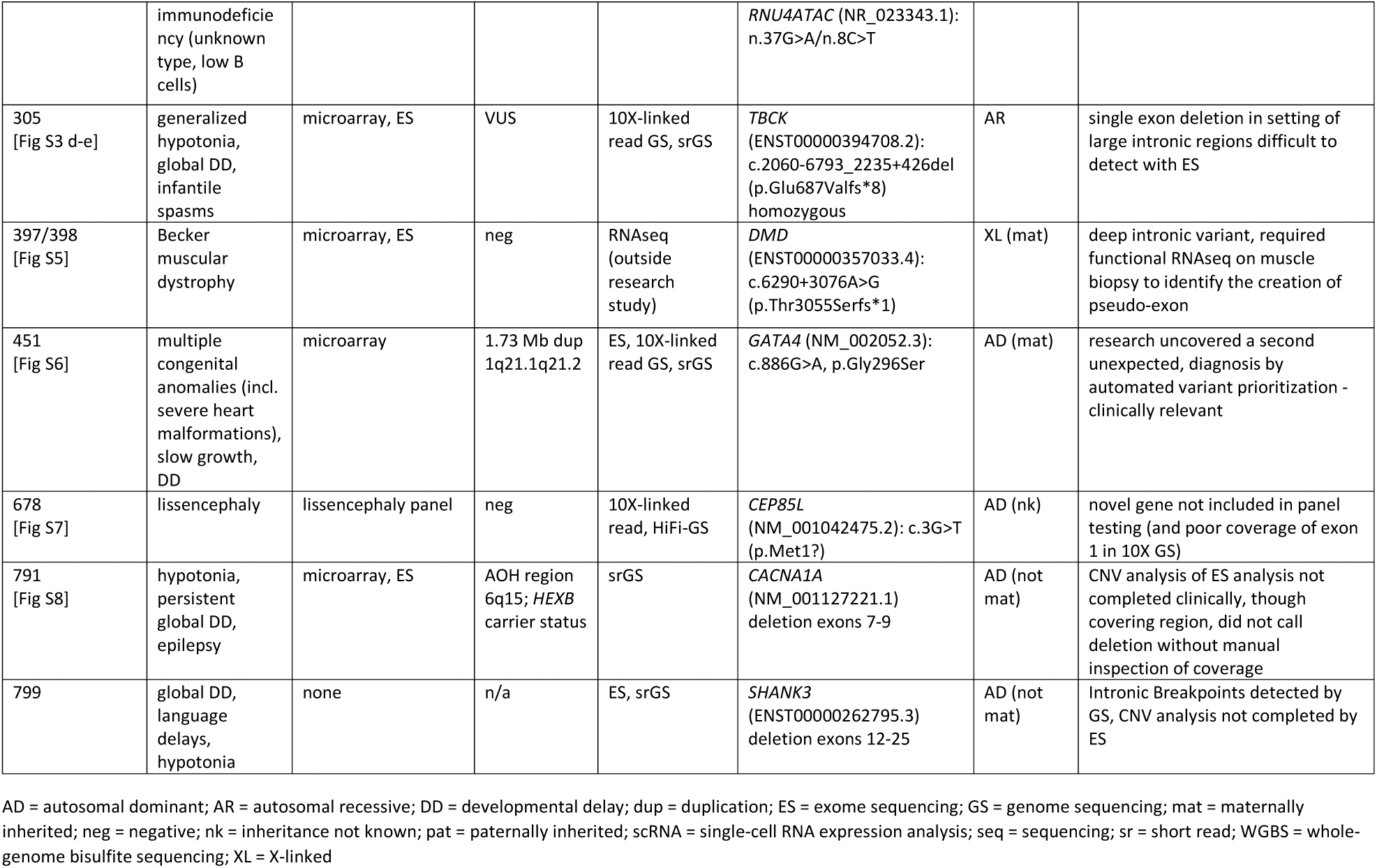
Example cases for which diagnosis was initially “missed” and subsequently solved through research analysis.

### Impact of GS platforms

GS contributed 6-13% of diagnoses (see above). The different platforms assessed in our study exhibited distinct characteristics that can contribute to individual variant types and overall potential for augmentation of ES. We examined three srGS platforms: 10X Linked sequencing (10X Genomics, n=587 total/542 patients), DNA NanoBall seq (MGI, n=180 total/74 patients) and PCR-free srGS (Illumina, n=1660 total/683 patients), along with a subset of samples assessed by HiFi-GS (PacBio, n=472 total/274 patients). The 10X Linked read sequencing exhibited inconsistent coverage across the genome, which resulted in suboptimal variant sensitivity (97.8% mean sensitivity), and we discontinued the method in favor of the other GS platforms which performed similarly (>98.3% sensitivity >98.8% specificity) against Infinium Global Screening microarray genotypes (Supplementary Figure S9). Considering the moderate increase in diagnostic yield with srGS, long-read genome sequencing utilizing PacBio HiFi reads was systematically deployed, allowing for a thorough comparison of HiFi-GS to srGS with a particular focus on the potential for rare disease variant discovery. Direct comparison of overall SNV calls and SV calls indicated an approximately 5% increase in SNV called from high coverage lrGS (25x HiFi WGS) vs. srGS (35x Illumina WGS), with a much more dramatic impact on SV detection with nearly double the discovery rate with lrGS (Supplementary Table S4).

To gauge the impact on potential rare disease SNV alleles, we compared a subset of probands (n=102) with both srWGS and HiFi-GS, focusing on rare coding variants. On average there were 476 coding variants per proband genome, of which 14% were unique to HiFi-GS, in contrast to 6% unique rare coding variants in srGS. Of these variants, transmission (variant detected in parent) supported nearly all (98%) variants observed by both srGS and HiFi-GS, whereas 40% of HiFi-GS specific variants appeared transmitted and 20% of srGS specific variants showed evidence of transmission. Extrapolating true positive rates per genome and per technology based on transmission suggests that on average lrGS exclusively detects 31 coding variants and srGS six coding variants per genome (Supplementary Table S5). More striking differences are observed for family-transmitted rare SVs (MAF <1%) generated at our center in either srGS or HiFi-GS data and not seen in publicly available reference data including DGV for srGS, HPRC HiFi-GS, or variants published from ONT-lrGS by Decode for lrGS.^19,34,35^ On average, 70 rare transmitted SVs are observed in srGS data and >300 for HiFi-GS: a greater than four-fold difference. The discovery advantage for HiFi-GS also applies for transmitted rare coding SVs (Table 2). Similarly to earlier reports, the rate of *de novo* SVs is low and only two (non-coding) examples were found in manual curation of eight high coverage HiFi-GS trios (Supplementary Table S6).^36^

**Table 2.** Structural variation.

|  | Average proband counts - Illumina/MGI srGS (49 trios), >30x Coverage |  |  |  |  |  |  |
| --- | --- | --- | --- | --- | --- | --- | --- |
|  | TOT | BND | CNV | DEL | DUP | INS | INV |
| All | 11036 | 2046 |  | 4299 | 393 | 4299 |  |
| Rare | 260 | 98 |  | 43 | 10 | 108 |  |
| Family-validated | 9127 | 1537 |  | 3876 | 339 | 3375 |  |
| Rare family-validated | 69 | 24 |  | 19 | 4 | 22 |  |
| Rare family-validated coding | 20 | 6 |  | 6 | 1 | 7 |  |

|  | Average proband counts – PacBio HiFi-GS (81 trios) > 25x (proband) >10x (parents) |  |  |  |  |  |  |
| --- | --- | --- | --- | --- | --- | --- | --- |
|  | TOT | BND | CNV | DEL | DUP | INS | INV |
| All | 22013 | 52 | 5 | 9104 | 412 | 12354 | 86 |
| Rare | 398 | 4 | 1 | 160 | 15 | 217 | 2 |
| Family-validated | 21114 | 45 | 5 | 8768 | 390 | 11824 | 81 |
| Rare family-validated | 332 | 3 | 1 | 136 | 12 | 179 | 2 |
| Rare family-validated coding | 119 | 1 | 0 | 46 | 4 | 67 | 1 |

### Enabling rare disease allele discovery by HiFi-GS

One tangible consequence of higher discovery rate of variant detection by HiFi-GS was the detection of 4,369,149 recurrent (observed in at least 2 unrelated individuals) SNVs not reported in gnomAD, as well as 115,595 recurrent SVs detected in our aggregated HiFi-GS resource (30,707 not seen in any previously published dataset).^37^ These findings serve as a reminder that publicly available datasets remain highly incomplete. To enable new rare disease discovery efforts by HiFi-GS, we are sharing these recurrent variants and their frequencies derived from >1,000 alleles of HiFi-GS data (https://github.com/ChildrensMercyResearchInstitute/GA4K). As anticipated, the recurrent variants detected in HiFi-GS were biased to regions with poor srGS resolution (e.g. segmental duplications and satellite repeats), but recurrent SVs not in DGV were widely dispersed across genic regions, and >800 OMIM loci also showed higher than GENCODE average rate of HiFi-GS specific SNVs (Supplementary Tables S7, S8).

The current diagnostic evaluation for rare disease relies on a multitude of genome-wide tests (ES, GS, microarray, chromosomes) as well as specialized directed tests (for repeat expansions, methylation defects, etc). We explored the potential for HiFi-GS to consolidate some of this testing and therefore reduce costs in the diagnostic odyssey for each proband. Developing a toolkit for HiFi-GS in rare disease included the accommodation of specific queries for known repeat expansion loci (Supplementary Table S9). Among our cohort, where each sample had a minimum 8x HiFi-GS coverage across 51 loci, we identified three pathogenic events (one *FMR1*, not shown, and two *STARD7* expansions, illustrated in Figure 3a-c). Additionally, while not specifically explored, there are known disease genes among the loci with an excess of “non-gnomad” variation (see above) such as *OTOA* and *STRC* which are challenging to test due to known pseudogenes/duplications (Supplementary Table S7). We note that the current alignment/variant calling pipeline for HiFi-GS also generates phased haplotypes, that allow detection of compound heterozygotes even in the case of singletons (Figure 3d-g), with an average phase block of 400kb (Supplementary Figure S10). Finally, the combination of SV calls and personal assemblies allowed the identification of HiFi-GS signatures for large CNVs clinically detected by microarray (Supplementary Table S10). Further, the implementation of personal assembly data can add basepair-level resolution for complex rearrangements interpreted as “balanced” by cytogenetic assays due to resolution limitations (Supplementary Figure S11).

**Figure 3.**
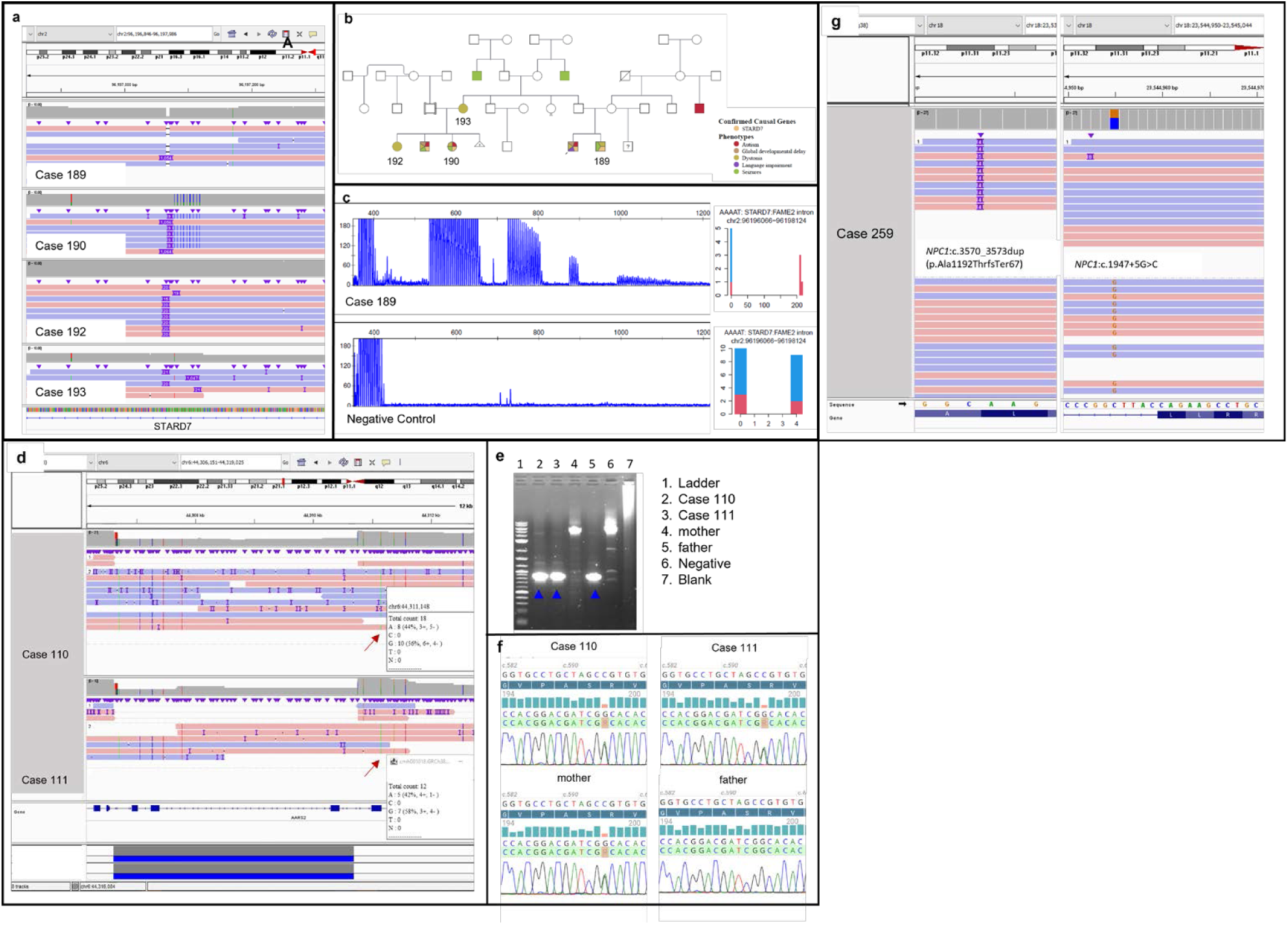
Examples of cases solved by HiFi-GS. Long read genome sequencing addresses challenges in srGS as exemplified by three cases. (**a**) HiFi-GS identified a novel pentamer expansion in *STARD7*, previously associated with Familial adult myoclonic epilepsy, 2 in an extended family. (**b**) Pedigree of family with *STARD7* disease, case 192 had adult-onset dystonia, while case 160 and case 189 had childhood onset of disease, consistent with anticipation. (**c**) Repeat primed-PCR confirmed the expansion detected in the HiFi-GS in case 189, which was also detected by the tandem genotyping tool. The negative control had a normal repeat pattern. (**d**) Affected siblings case 110 and case 111 were found to be compound heterozygous for two pathogenic variants in *AARS2*, NM_020745.4: c.595C>T (p.Arg199Cys), maternally inherited, and a paternally inherited deletion, chr6:44306625-44310745 encompassing exons 5-7 of *AARS2*. (**e**) Clinical confirmation of the deletion using long read PCR detected the deletion (arrow) and normal allele in both siblings and unaffected father. (**f**) Clinical sanger confirmation of the maternally inherited c.595C>T (p.Arg199Cys) variant. (**g**) case 259 was clinically diagnosed with Niemann-Pick disease, but parents were unavailable for phasing. HiFi-GS confirmed the pathogenic variants were *in trans*, consistent with autosomal recessive disease. *NPC1*: c.3570_3573dupACTT (p.Ala1192Thrfs*67) (left)/ c.1947+5G>C (right).

### New candidate genes following re-analysis across all data and variant prioritization

The joint sequencing results and automated prioritization were reviewed by an expert analysist (genetic counsellor or clinical laboratory director) to identify a large fraction of patients (58%) with potential new disease genes. Compelling candidates were systematically submitted to GeneMatcher (GM).^5^ At the time of manuscript submission, 152 candidate genes were active in GM, 12 of which were identified in more than one unrelated family, and six of which were recently published or close to publication and therefore in transition from GUS to diagnostic. More than 36% of submitted GUS had more than 10 hits in GM, suggesting they are strong candidates. This underscores the imperative for data sharing and collaboration in rare disease research and diagnosis.

### Individual data sharing to enhance variant and gene discovery

Uniform research consents permit sharing of sequences and structured phenotypic data with other rare disease investigators to enhance gene matching beyond the variation submitted to GeneMatcher. Raw data submitted to dbGAP (phs002206.v2.p1) will allow for joint calling with other available rare disease datasets. Access to processed data for rare variants, de-identified pedigrees and coded phenotypes will be available to registered users through a cloud-hosted PhenoTips web UI: https://phenotips-ga4k.cmh.edu (access inquiries for investigators) (Supplementary Figure S12).^24^ This web UI provides a simple interface for users to review participant data, identify cohorts of participants based on phenotypic or genotypic attributes, and review rare variants in the context of a specific phenotype. Furthermore, this interface will continue to be dynamically synchronized with the GA4K program, and already included >1000 additional cases in various stages of ongoing analyses at the time of manuscript submission, for a total of 5922 individuals across 2537 families and processed variants for 2069 patients.

## DISCUSSION

We developed a comprehensive rare disease phenotype-genotype data repository across a large pediatric healthcare system in the Genomic Answers for Kids program. Full access is provided to enable medical genomic testing, complete annotation for reanalysis, and use by contemporary research genomic tools. Using multiple sequencing methods and analytic approaches the first 1,083 patients evaluated serve as a roadmap to improve rare disease diagnostics, and as a catalog of case data for utility in biomedical discovery.

We combined publicly available machine learning approaches Exomiser and AMELIE (“E/A”) for variant and disease gene prioritization at scale where the nominated candidate variant was ranked by E/A in 539 (49.8%) cases, supporting the use of machine-learning tools as a first-pass, resource-saving analysis. Primarily retrospective studies suggested higher rates of relevant results, and we replicate similar success to these studies in our observed concordance among previously diagnosed cases.^7,30,38^ Importantly, the vast majority of our patients were undiagnosed when entering the study. This allowed us to establish the utility of computationally assisted interpretation among prospective diverse rare disease patients, on a scale far beyond any previously assessed rare disease cohort.^9,38^ We also showcase patients having a strong candidate or diagnostic variant identified through machine-learning ranking (subsequently confirmed through expert review) that may not have otherwise been prioritized for further investigation due to combined supporting data being pulled by artificial intelligence from multiple sources and not easily digested by manual analysis in a timely manner, as expected in a clinical setting (i.e. not an obvious candidate that would arise from easily checked metrics such as gene constraint and protein function). This supports the utility of the approach, not only for diagnostic evaluation, but also as a systematic source for generating hypotheses on disease gene discovery. Importantly, prioritization is still biased given that it will inevitably rank genes that have more linked resources (be those clinical, functional, or otherwise) higher than poorly characterized genes, and therefore genetic prioritization independent of literature mining remains important for gene discovery.^39^

We demonstrated there is diagnostic utility in ES re-analyses and/or repeat ES to improve coverage; however, more than 10% diagnoses we made in previously negative ES cases were solved with elevation to GS which, unlike most ES analyses, included systematic CNV calling. As expected, the utility of GS was lower in previously unassessed cases, however even in this group 1/20 diagnoses required GS. Similarly to previously unsolved cases, GS contributed primarily to the detection of SVs. Given known benefits of HiFi-GS in SV detection, we pursued HiFi-GS in unsolved rare diseases beyond earlier demonstration studies as routine streamlining of trios.^19,20^ Early results from HiFi-GS demonstrated the expected improvement in detection rates for SVs, but also provided first glimpses of diagnostic variation currently only achievable by HiFi-GS such as the discovery of novel repeat expansions (including repeat size and sequence composition), the solving of CNV breakpoints and orientation/localization, and the resolution of phase in the absence of parental samples. The potential for having full genome analyses by HiFi-GS was explored here as proof-of concept; further work will elaborate underexplored areas of HiFi-GS utility, such as personal assemblies, haplotype-phasing, and directed work on duplicated gene regions. In the meantime, our HiFi-GS variant catalogs extending across hundreds of individuals provide the first building blocks for using alternative GS methods in clinical settings and particularly for unsolved diseases.

Finally, the majority of unsolved cases in our cohort do have candidate genes and variants but lack sufficient evidence to assign pathogenicity due to a lack of replication (also known as the “n of 1” problem), with hundreds of genes and variants currently followed through GeneMatcher. Greater data sharing is paramount for enhancing benefits to participants and advancing scientific progress, along with maximizing the utility of genomic data.^40^ Unfortunately, hesitancy towards extensive data sharing persists among investigators due to reasons that include the arduous processes required for data sharing, concerns about participant privacy, and fear for loss of priority in data publication.^40,41^ Our study follows regulations and considers recommendations for responsible sharing of pediatric genomic data to support the benefits of data sharing to research participants and patients while protecting privacy.^40^

## Data Availability

Processed data for rare variants, de-identified pedigrees and coded phenotypes available to registered users through a cloud-hosted PhenoTips web UI (https://phenotips-ga4k.cmh.edu/). Access inquiries for investigators should be directed to (including key to correlate study numbers used in this manuscript).
Recurrent variants and their frequencies derived from >1,000 alleles of HiFi-GS data available at: https://github.com/ChildrensMercyResearchInstitute/GA4K

https://phenotips-ga4k.cmh.edu/

## ACKNOWLEDGMENTS

We would like to thank the families for participating in our study. This work was made possible by the generous gifts to Children’s Mercy Research Institute and Genomic Answers for Kids program at Children’s Mercy Kansas City.

