## Supplemental Figures for "IGenomic answers for children: Dynamic analyses of >1000 pediatric rare disease genomes"

**Supplemental Figure S1.** Family of 239. (**a**) Pedigree illustrating the two siblings, who were referred for familial lipodystrophy; clinical exome sequencing was nondiagnostic. (**b**) Research reanalysis revealed a homozygous missense variant in *MFN2*: c.2119C>T (p.Arg707Trp), associated with multiple symmetric lipomatosis, representing a phenotypic expansion of the disease.

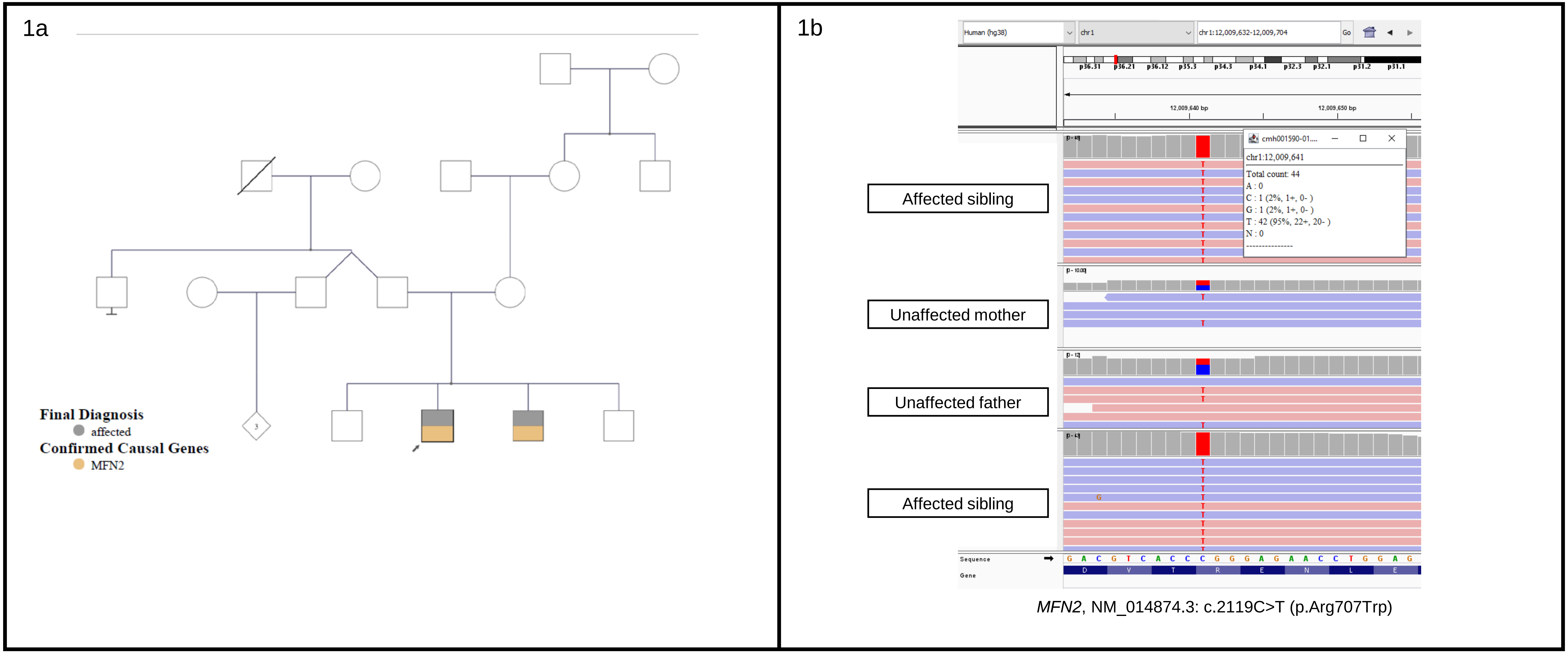

**Supplemental Figure S2.** Family of 453. (**a**) Pedigree illustrating the proband who was referred for myotonia congenita, and significant family history of disease. Clinical panel testing was negative. (**b**) Research testing revealed a known pathogenic variant in *SCN4A,* c.4342C>T (p.Arg1448Cys), associated with paramyotonia congenita. Follow-up communication revealed that the variant had not meet quality metrics in the original send-out testing and had not been reported, illustrating the importance of coverage and test-specific limitations stated by the laboratory.

2a

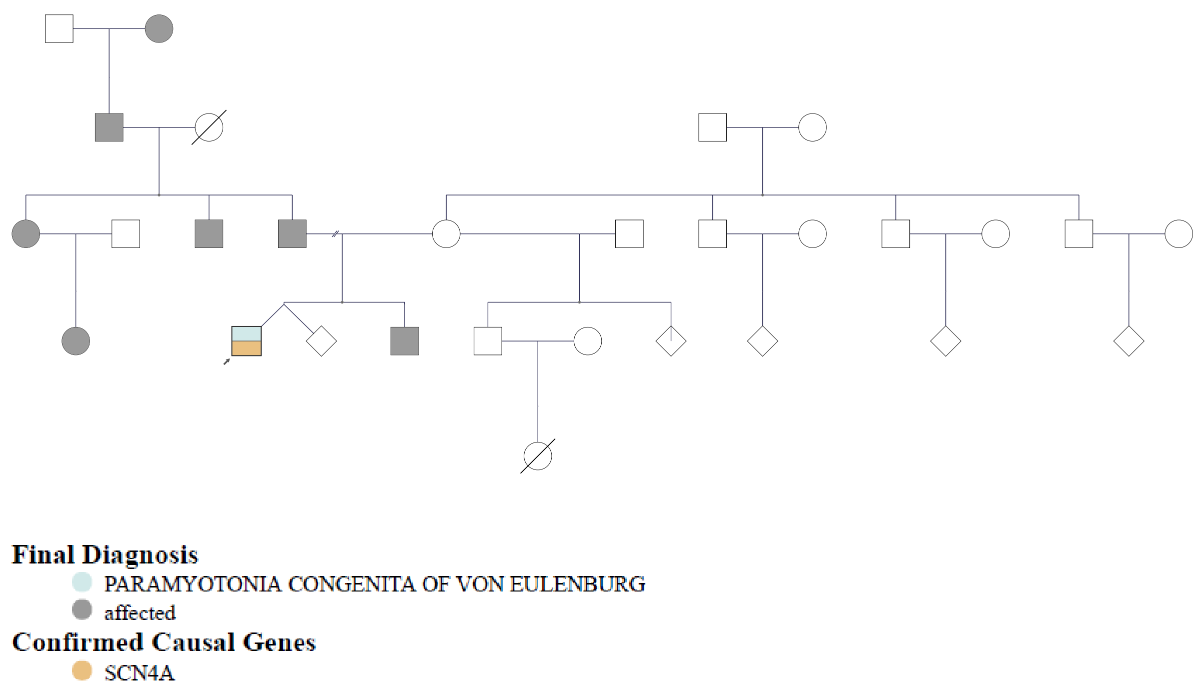

2b

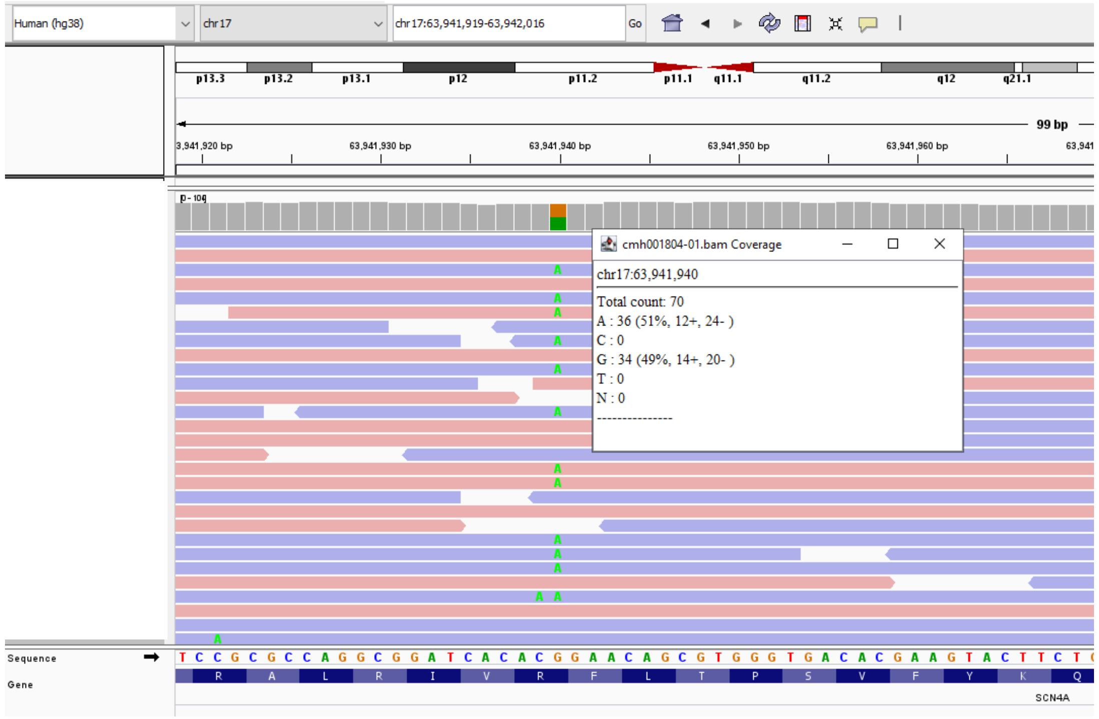

*SCN4A*, NM_000334.4: c.4342C>T (p.Arg1448Cys)

**Supplemental Figure S3.** Small copy number variants involving single exons that remain challenging to detect. (**a**) Pedigree of family of 129 proband who was referred for profound congenital hypotonia, with negative clinical testing. (**b**) Exome sequencing revealed a maternally inherited premature stop variant in *TBCK*:c.1039C>T(p.Arg347Ter), however a second variant was not identified. (**c**) Subsequent genome sequencing revealed a recurrent, paternally inherited deletion of exon 3 in *TBCK*: c.2060-6793_2235+426del (p.GLu687Valfs*8), consistent with a diagnosis of infantile hypotonia with psychomotor retardation and characteristic facies 3 (OMIM 616900). (**d**) Pedigree of family of 305 proband who was also referred for profound congenital hypotonia, with prior negative clinical testing. (**e**) Genome sequencing revealed homozygosity for the recurrent exon 23 deletion in *TBCK*, confirming the diagnosis of infantile hypotonia with psychomotor retardation and characteristic facies 3.

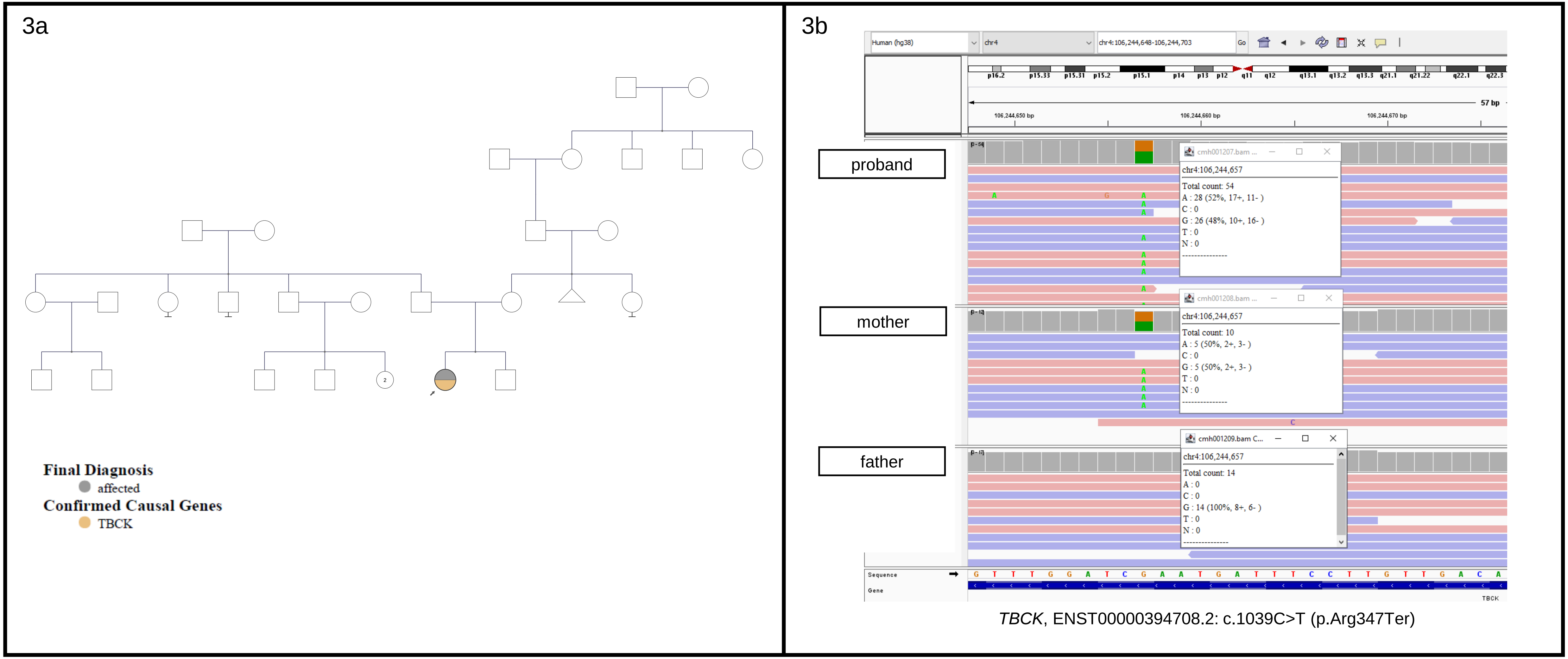

**Supplemental Figure S3.** Small copy number variants involving single exons that remain challenging to detect. (**a**) Pedigree of family of 129 proband who was referred for profound congenital hypotonia, with negative clinical testing. (**b**) Exome sequencing revealed a maternally inherited premature stop variant in *TBCK*:c.1039C>T(p.Arg347Ter), however a second variant was not identified. (**c**) Subsequent genome sequencing revealed a recurrent, paternally inherited deletion of exon 3 in *TBCK*: c.2060-6793_2235+426del (p.GLu687Valfs*8), consistent with a diagnosis of infantile hypotonia with psychomotor retardation and characteristic facies 3 (OMIM 616900). (**d**) Pedigree of family of 305 proband who was also referred for profound congenital hypotonia, with prior negative clinical testing. (**e**) Genome sequencing revealed homozygosity for the recurrent exon 23 deletion in *TBCK*, confirming the diagnosis of infantile hypotonia with psychomotor retardation and characteristic facies 3.

3c

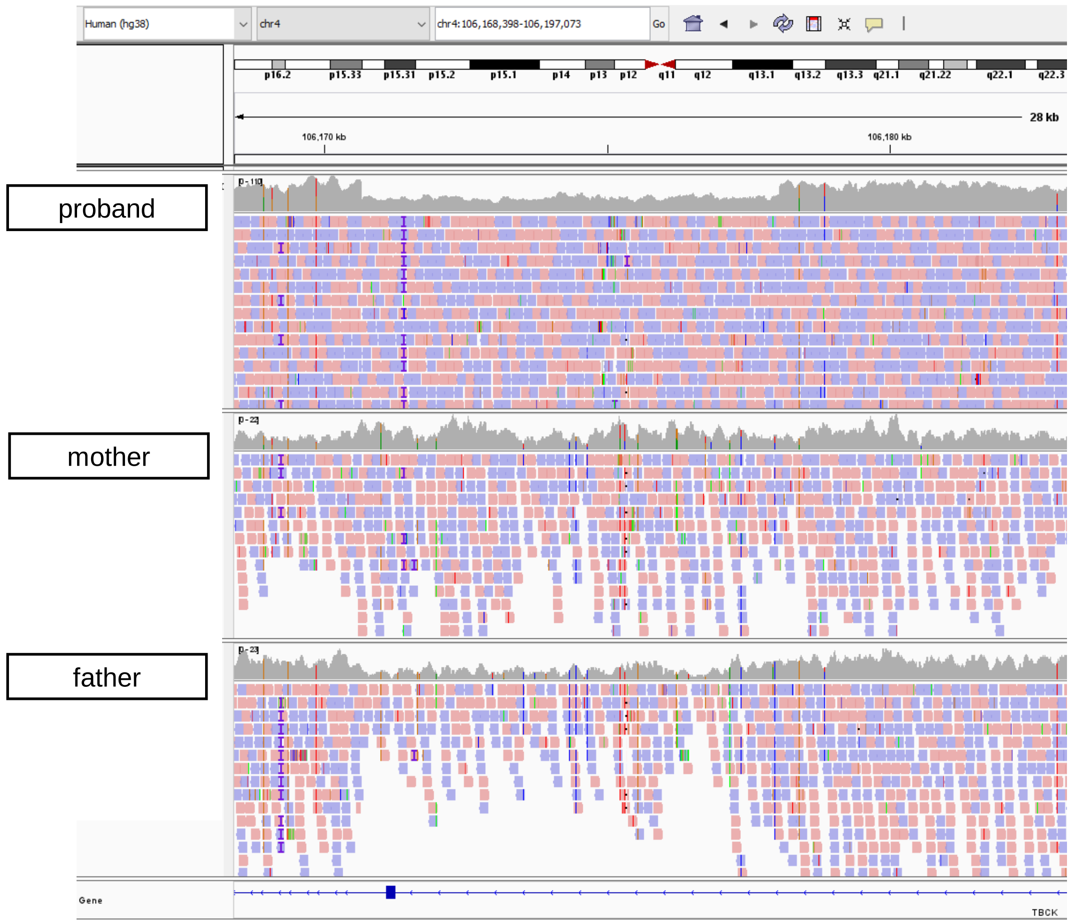

*TBCK*, ENST00000394708.2: c.2060-6793_2235+426del (p.Glu687Valfs*8)

**Supplemental Figure S3.** Small copy number variants involving single exons that remain challenging to detect. (**a**) Pedigree of family of 129 proband who was referred for profound congenital hypotonia, with negative clinical testing. (**b**) Exome sequencing revealed a maternally inherited premature stop variant in *TBCK*:c.1039C>T(p.Arg347Ter), however a second variant was not identified. (**c**) Subsequent genome sequencing revealed a recurrent, paternally inherited deletion of exon 3 in *TBCK*: c.2060-6793_2235+426del (p.GLu687Valfs*8), consistent with a diagnosis of infantile hypotonia with psychomotor retardation and characteristic facies 3 (OMIM 616900). (**d**) Pedigree of family of 305 proband who was also referred for profound congenital hypotonia, with prior negative clinical testing. (**e**) Genome sequencing revealed homozygosity for the recurrent exon 23 deletion in *TBCK*, confirming the diagnosis of infantile hypotonia with psychomotor retardation and characteristic facies 3.

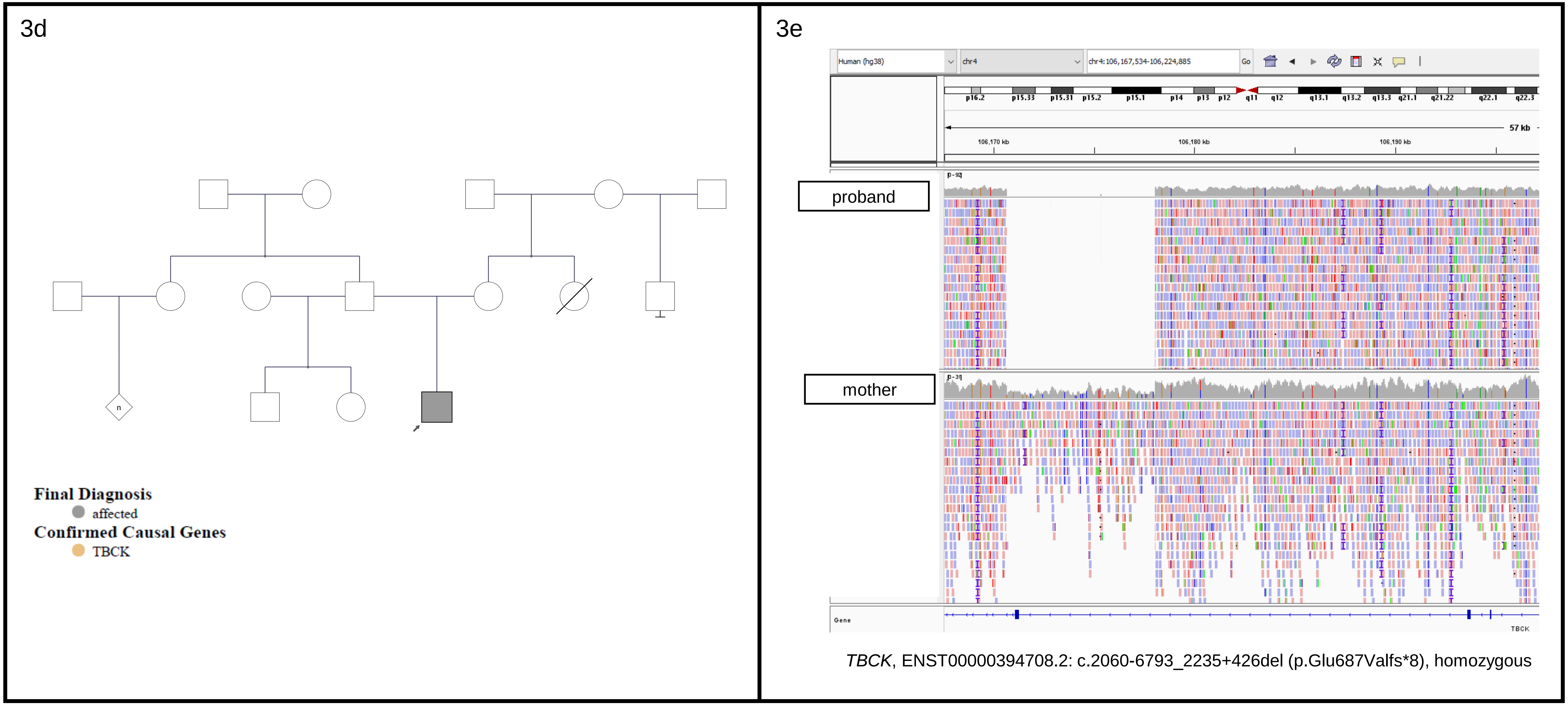

*TBCK*, ENST00000394708.2: c.2060-6793_2235+426del (p.Glu687Valfs*8), homozygous

**Supplemental Figure S4**: Family of 302. (**a**) Pedigree of proband who was referred due to multiple affected siblings with immunodeficiency, small growth, small hands and feet, and variable developmental delays. (**b**) Clinical testing revealed a paternally-inherited premature stop variant in *SPECC1L*: c.1900C>T (p.Arg634Ter), which did not explain the full phenotype of the siblings (OMIM 614140). (**c**) Subsequent genome sequencing revealed a second diagnosis of Roiffman syndome, an autosomal recessive disorder caused by variants in *RNU4ATAC* which encodes a non-coding RNA not covered by exome testing.

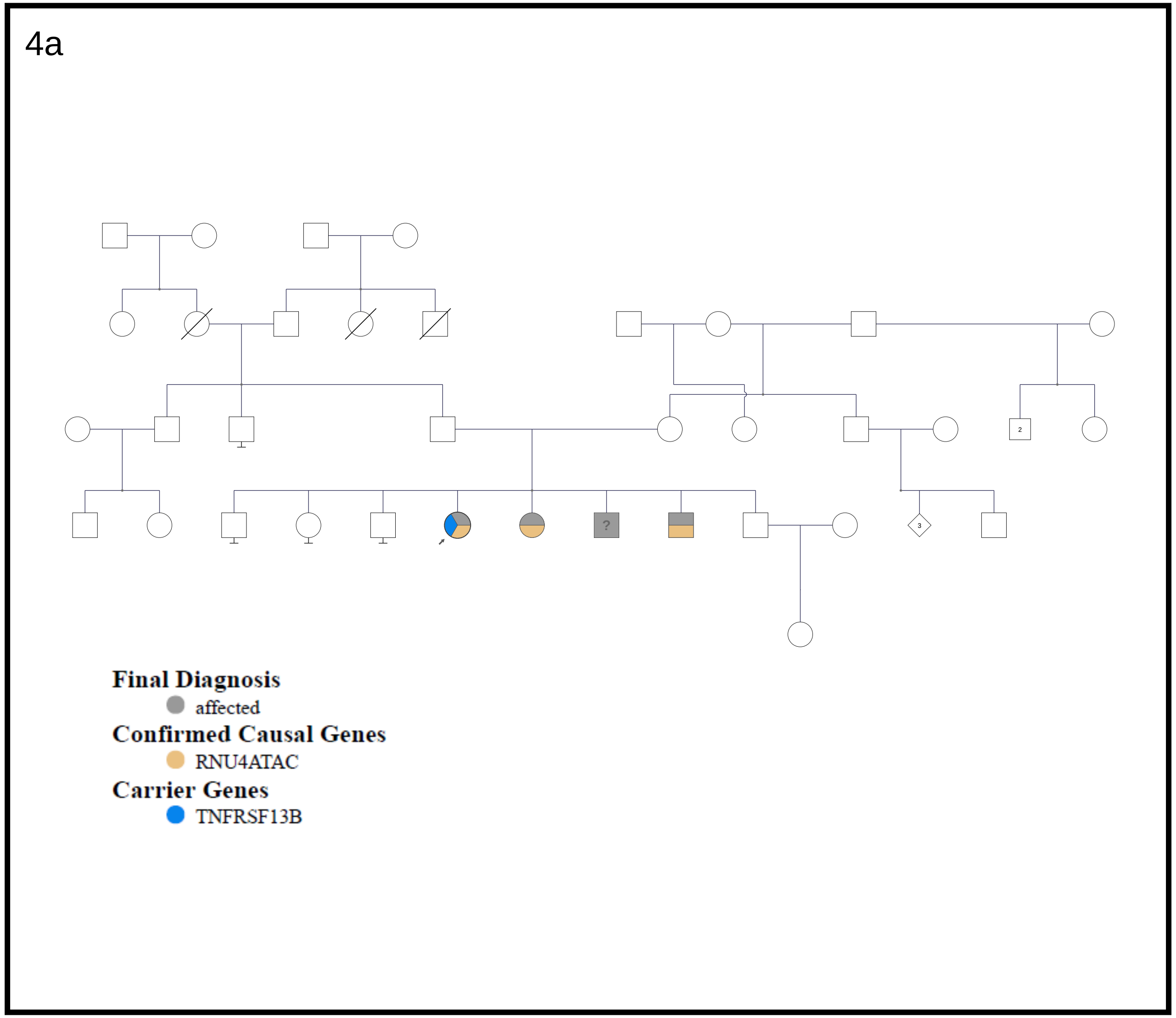

**Supplemental Figure S4**: Family of 302. (**a**) Pedigree of proband who was referred due to multiple affected siblings with immunodeficiency, small growth, small hands and feet, and variable developmental delays. (**b**) Clinical testing revealed a paternally-inherited premature stop variant in *SPECC1L*: c.1900C>T (p.Arg634Ter), which did not explain the full phenotype of the siblings (OMIM 614140). (**c**) Subsequent genome sequencing revealed a second diagnosis of Roiffman syndome, an autosomal recessive disorder caused by variants in *RNU4ATAC* which encodes a non-coding RNA not covered by exome testing.

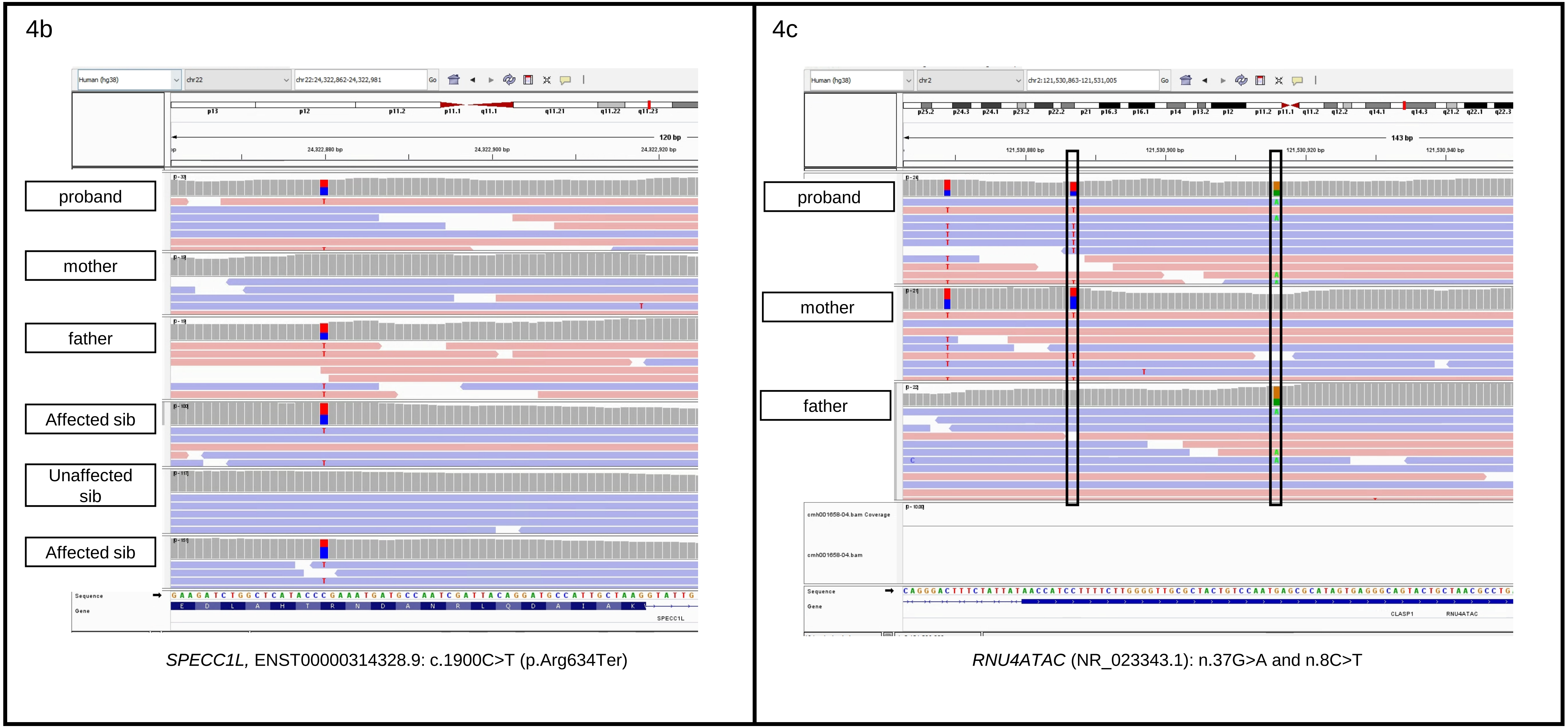

*SPECC1L,* ENST00000314328.9: c.1900C>T (p.Arg634Ter)

*RNU4ATAC* (NR_023343.1): n.37G>A and n.8C>T

**Supplemental Figure S5.** Family of 397. (**a**) Pedigree illustrating the proband and his male sibling, who both had a clinical presentation of Becker muscular dystrophy (**b**) Previously completed research testing had identified a deep intronic variant in *DMD*, only covered by genome sequencing. Additional functional studies (data not shown) demonstrated that the variant creates an alternative splice site (predicted to result in inclusion of a pseudoexon 43), leading to disease.

5a

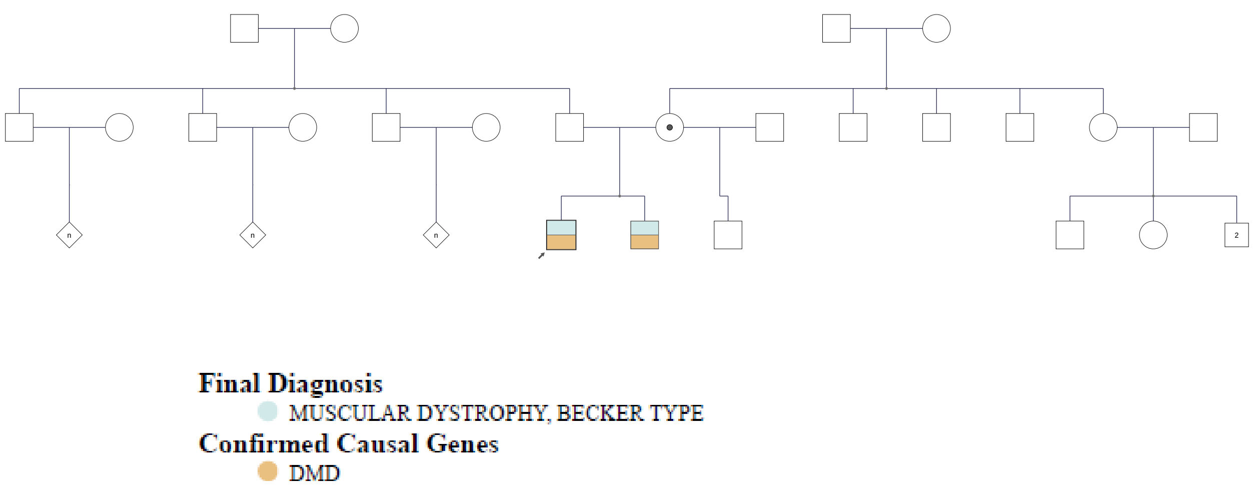

5b

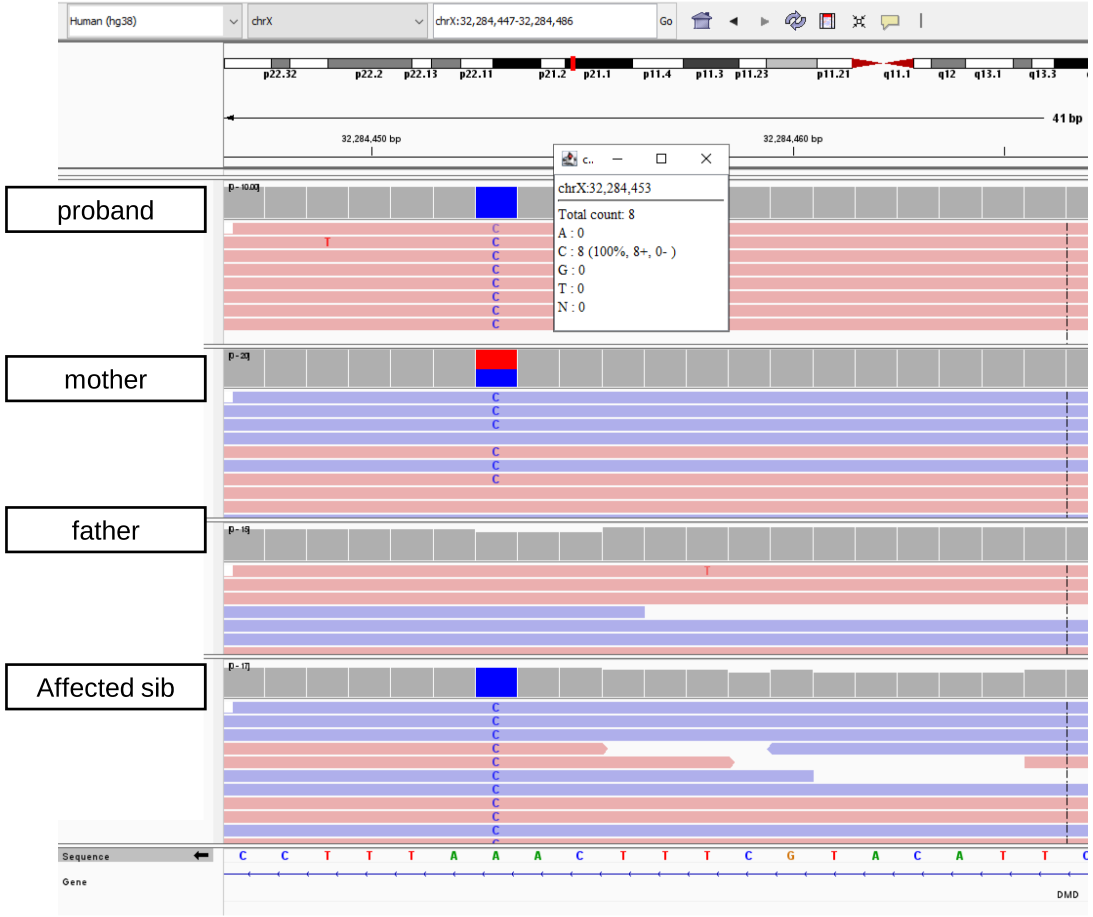

*DMD,* ENST00000357033.4: c.6290+3076 A>G

**Supplemental Figure S6.** Family of 451. (**a**) Pedigree illustrating proband who was diagnosed prenatally due to multiple anomalies with a maternally-inherited pathogenic duplication of 1.73 Mb at 1q21.1q21.2. (**b**) Subsequent research analysis (and E/A prioritization) identified a second diagnostic finding: a maternally-inherited pathogenic variant in *GATA4*: c.889G>A (p.Gly297Ser), associated with congenital heart defects

6a

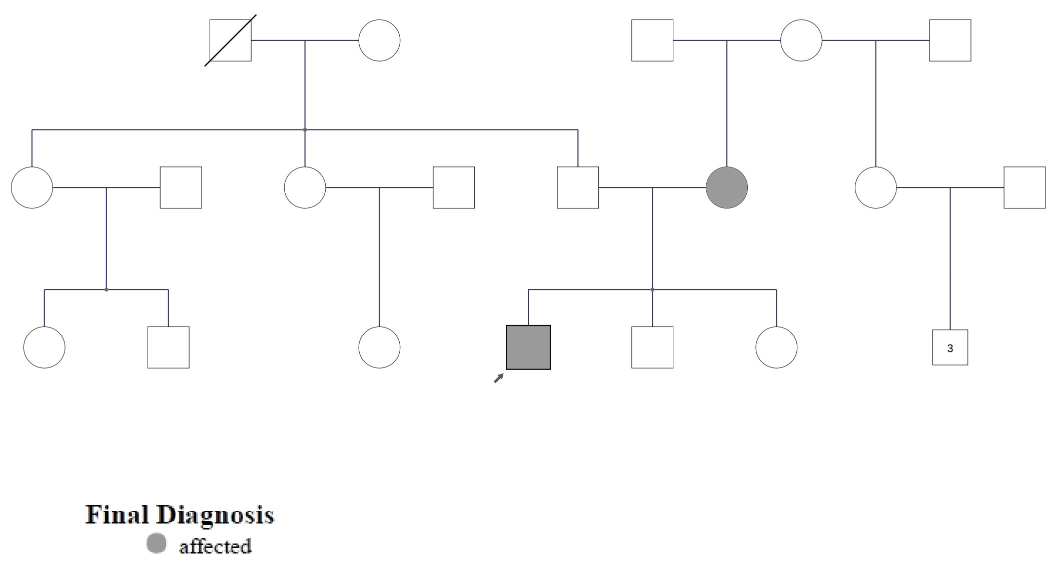

Dual Diagnosis: arr[GRCh37] 1q21.1q21.2(146137165_147867742)x3 mat

6b

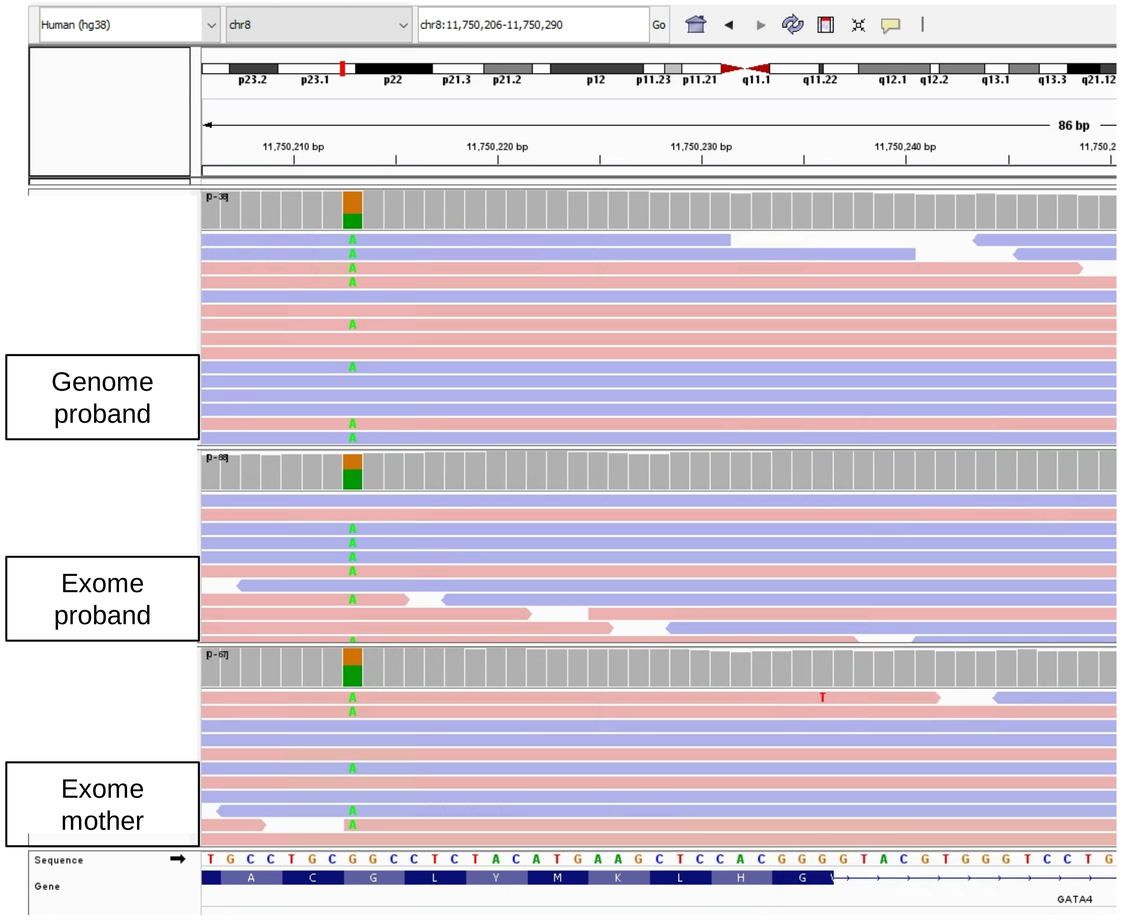

*GATA4*, NM_002052.3: c.889G>A (p.Gly297Ser)

**Supplemental Figure S7.** Family of 678. (**a**) Pedigree of family 678 who was referred for lissencephaly and mild intellectual disability with a negative family history. (**b**) Initial testing using 10X linked read genome sequencing was non-diagnostic; however, subsequent sequencing using PacBio HiFi reads revealed a heterozygous known pathogenic variant in *CEP85L*: c.3G>T (p.Met1?), associated with autosomal dominant lissencephaly.

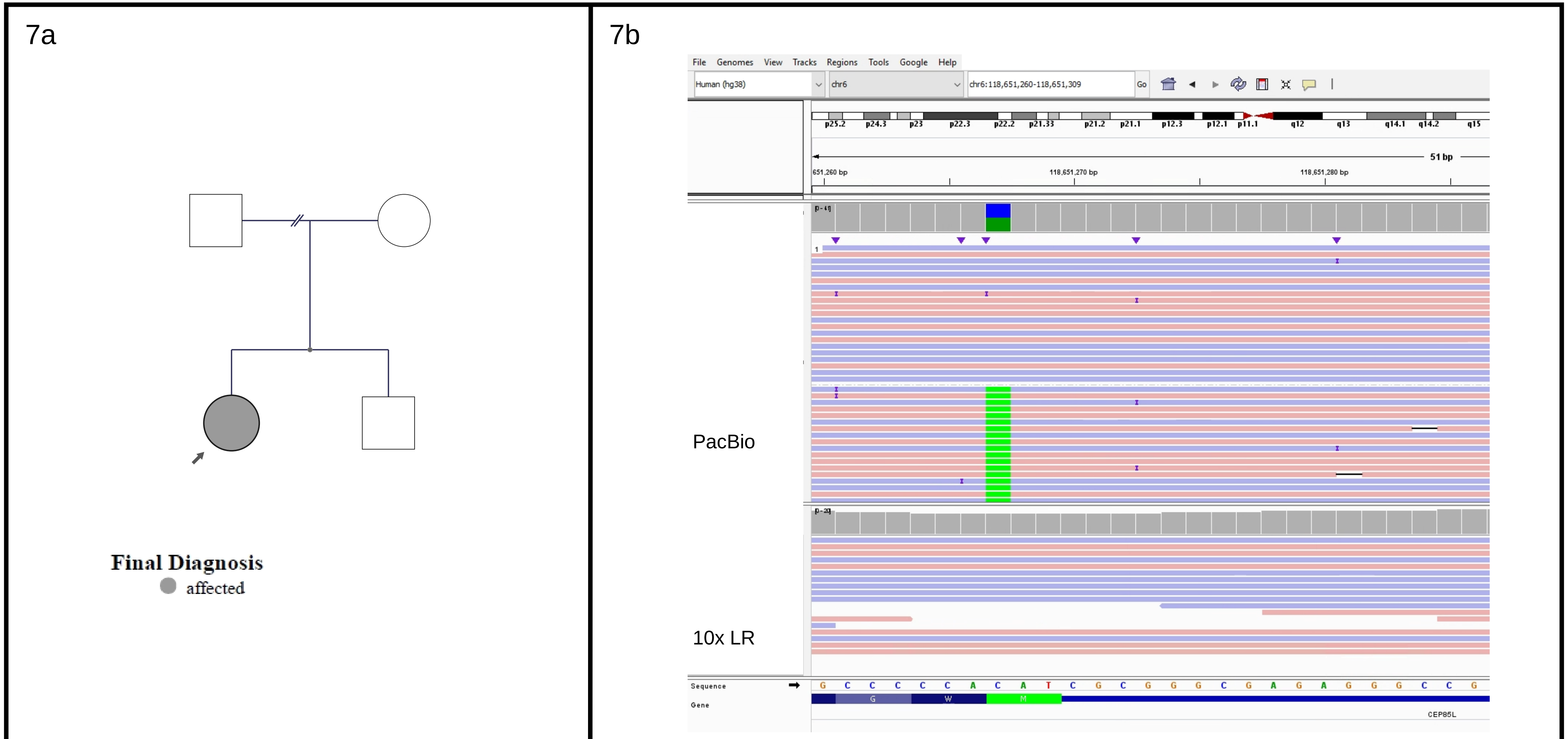

*CEP85L*, NM_001042475.2: c.3G>T (p.Met1?)

**Supplemental Figure S8.** Family of 791. (**a**) Pedigree of proband who was referred for failure to thrive, hypotonia, persistent global developmental delays, and epilepsy, with a negative family history. (**b**) Genome sequencing revealed a deletion of exons 7-8 and part of exon 9 of *CACNA1A*: c.979-354_1224del, predicted to result in a frameshift and premature stop, consistent with a diagnosis of developmental and epileptic encephalopathy 42 (OMIM 617106).

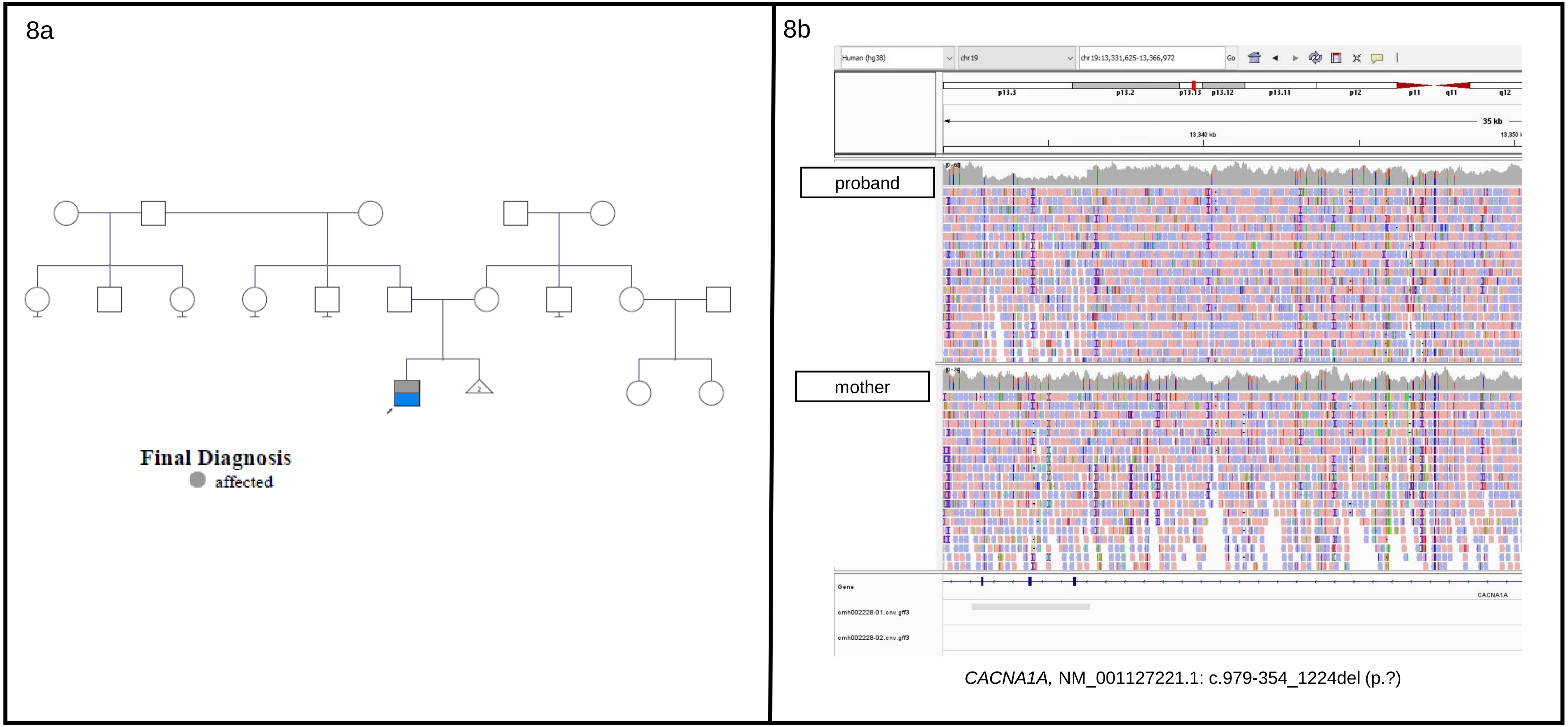

*CACNA1A,* NM_001127221.1: c.979-354_1224del (p.?)

**Supplemental Figure S9.** (**a**) Sensitivity of SNV calls by technology compared to a truth set of calls from an Illumina genotypic array. 10X Linked read GS consistently had lower sensitivity, although still greater than 95%. (**b**) Specificity of SNV calls by technology compared to a truth set of calls from an Illumina genotypic array.

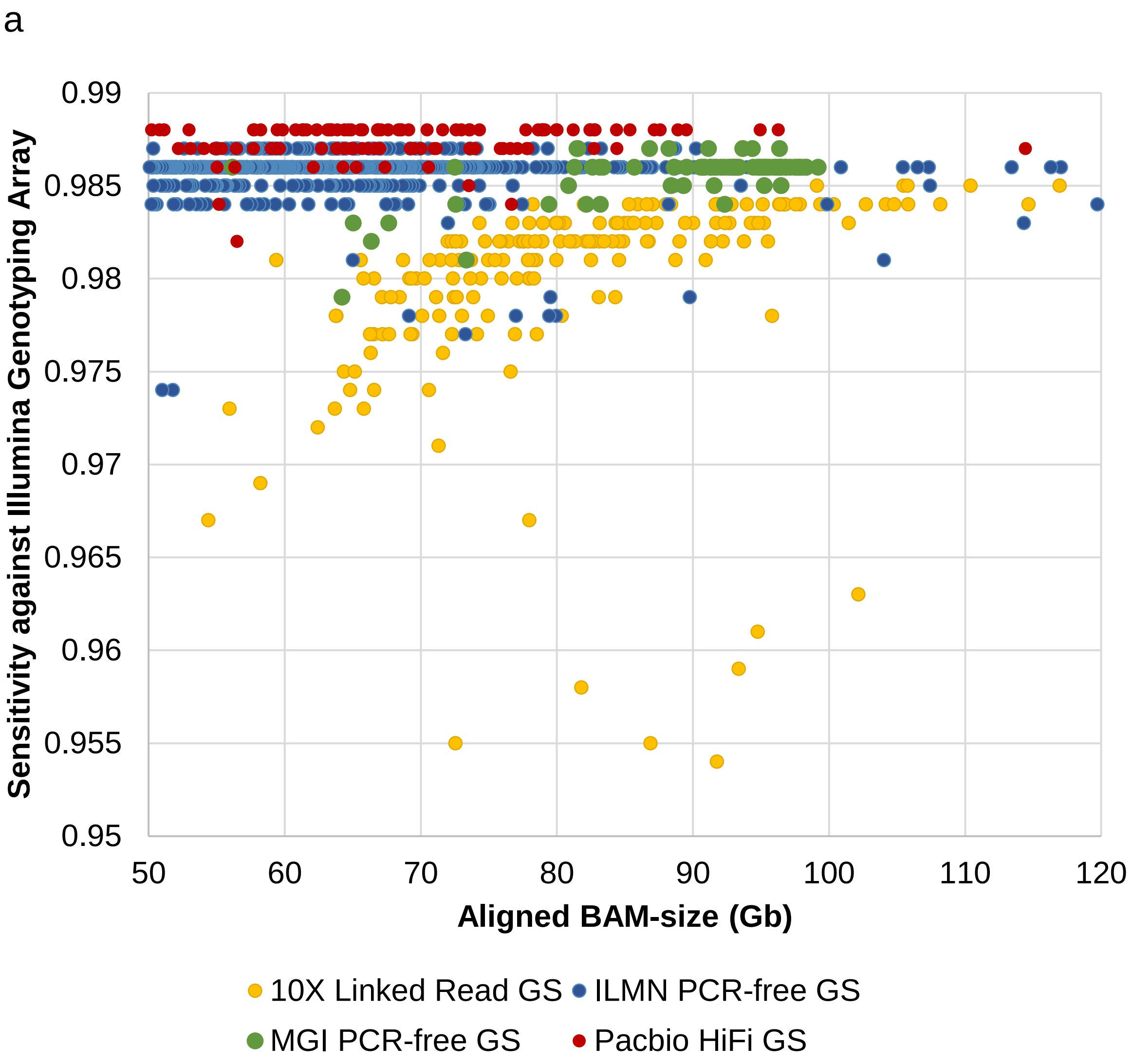

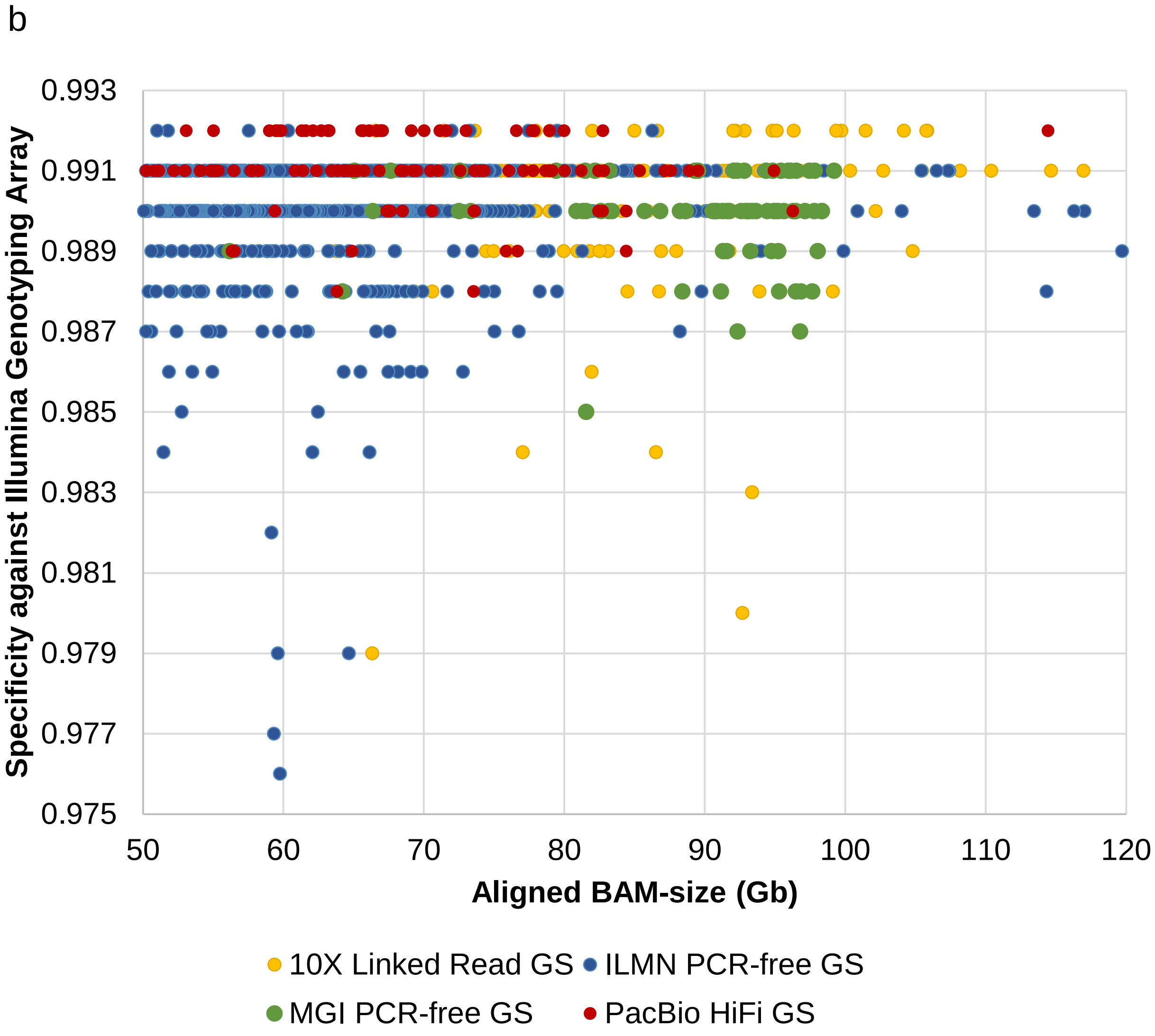

**Supplemental Figure S10.** The average N50 phase block generated from HiFi-GS is 400kb in the first 80 HiFi-GS samples. This can provide accurate phasing information, which is particularly important for patients where parental samples are not available.

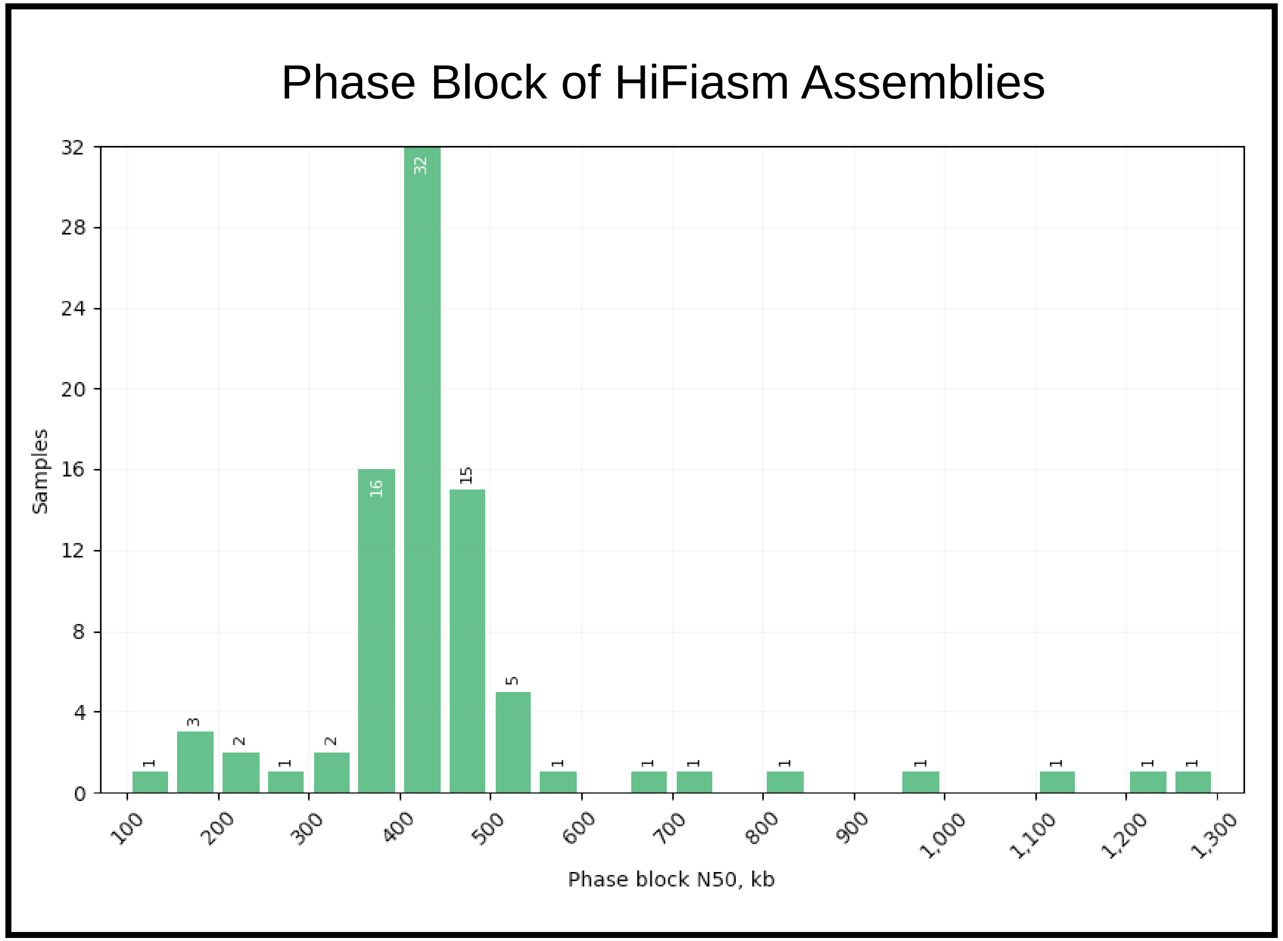

**Supplementary Figure S11.** Cytogenetically-detected apparently balanced translocation, shown to lead to net loss of 1.3Mb in chr2 by PacBio HiFi-GS with a complex structural rearrangement. (**a**) Breakends detected by HiFi-GS (**b**) Graphical representation of structural variation using Ribbon (**c**) Genomic content impacted by SV

11a

| | | | | | | |
|---|---|---|---|---|---|---|
| ptg000019l | 21 | 303894 | chr2 | 15840701 | 16144748 | - |
| ptg000019l | 303925 | 19379010 | chr9 | 75260460 | 94312900 | + |
| ptg000049l | 391 | 7102660 | chr2 | 7489537 | 14582268 | + |
| ptg000049l | 7102670 | 8393768 | chr2 | 67161593 | 68451729 | + |
| ptg000148l | 8 | 694836 | chr9 | 68220556 | 68915363 | - |
| ptg000148l | 695521 | 726211 | chr9 | 63809315 | 63838794 | - |
| ptg000148l | 730706 | 866472 | chr9 | 62822795 | 62958343 | + |
| ptg000148l | 745559 | 769050 | chr9 | 63759161 | 63782370 | - |
| ptg000148l | 853425 | 913874 | chr9 | 62822795 | 62883135 | - |
| ptg000148l | 918369 | 949078 | chr9 | 63809315 | 63838794 | + |
| ptg000148l | 949763 | 3443669 | chr9 | 68220556 | 70719798 | + |
| ptg000148l | 3425660 | 7971287 | chr9 | 70719799 | 75260436 | + |
| ptg000148l | 7971287 | 9230075 | chr2 | 14582291 | 15840687 | + |
| ptg000148l | 9230087 | 9371741 | chr2 | 67019935 | 67161573 | - |

11b

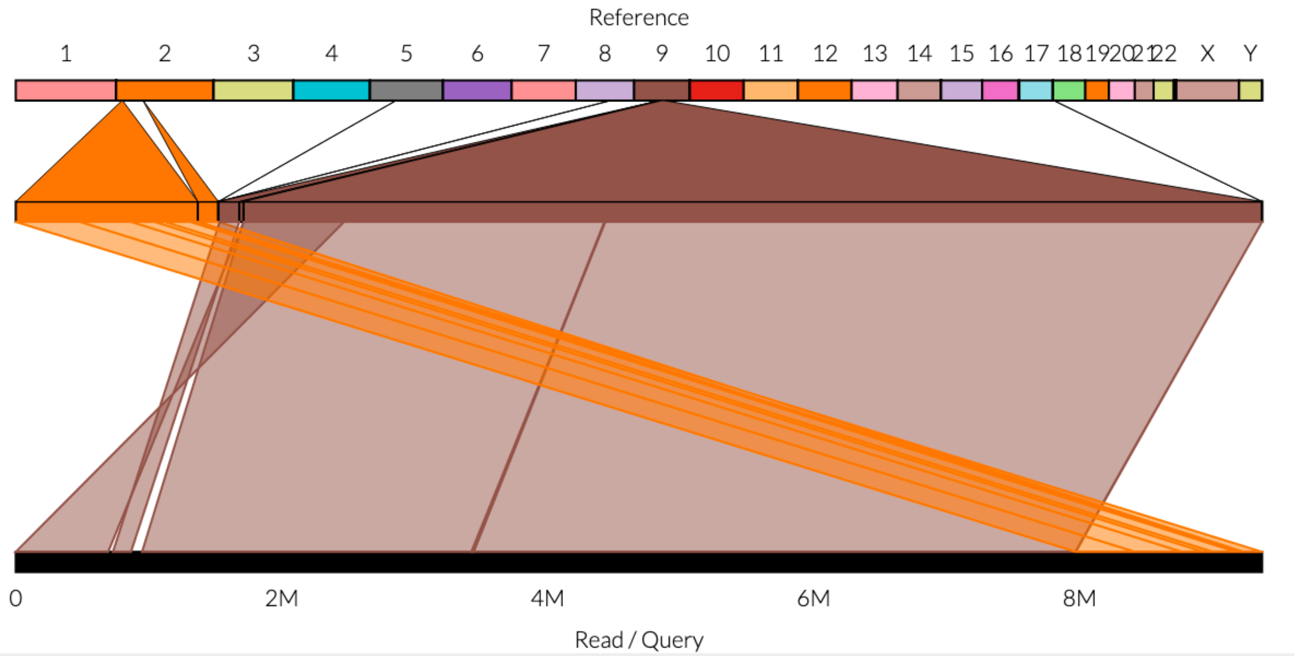

11c

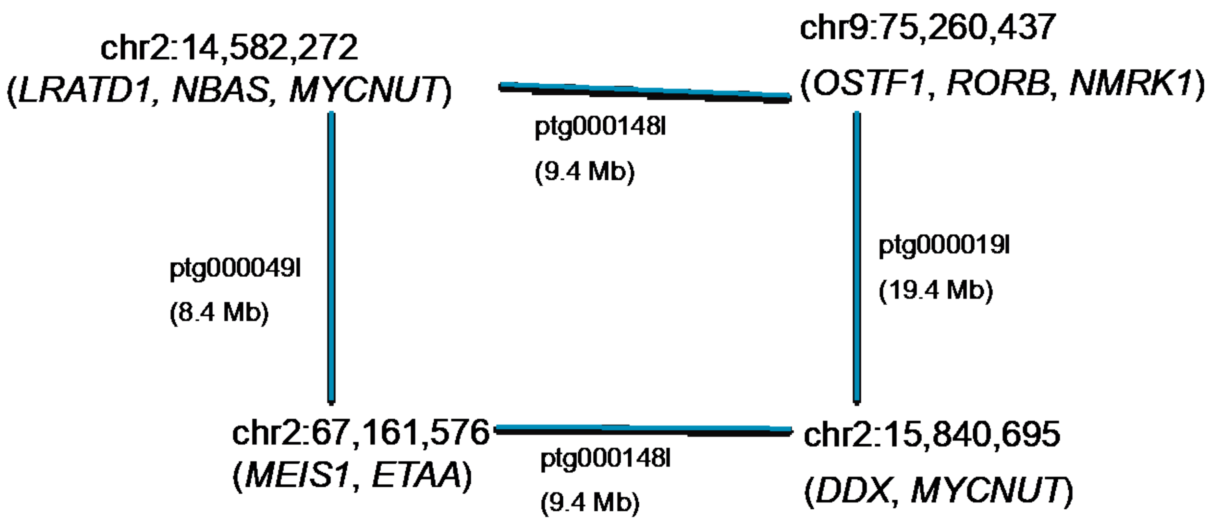

**Supplemental Figure S12.** Screenshot to illustrate how to navigate publicly accessibly data, described in steps a to g. This is available at https://phenotips-ga4k.cmh.edu/

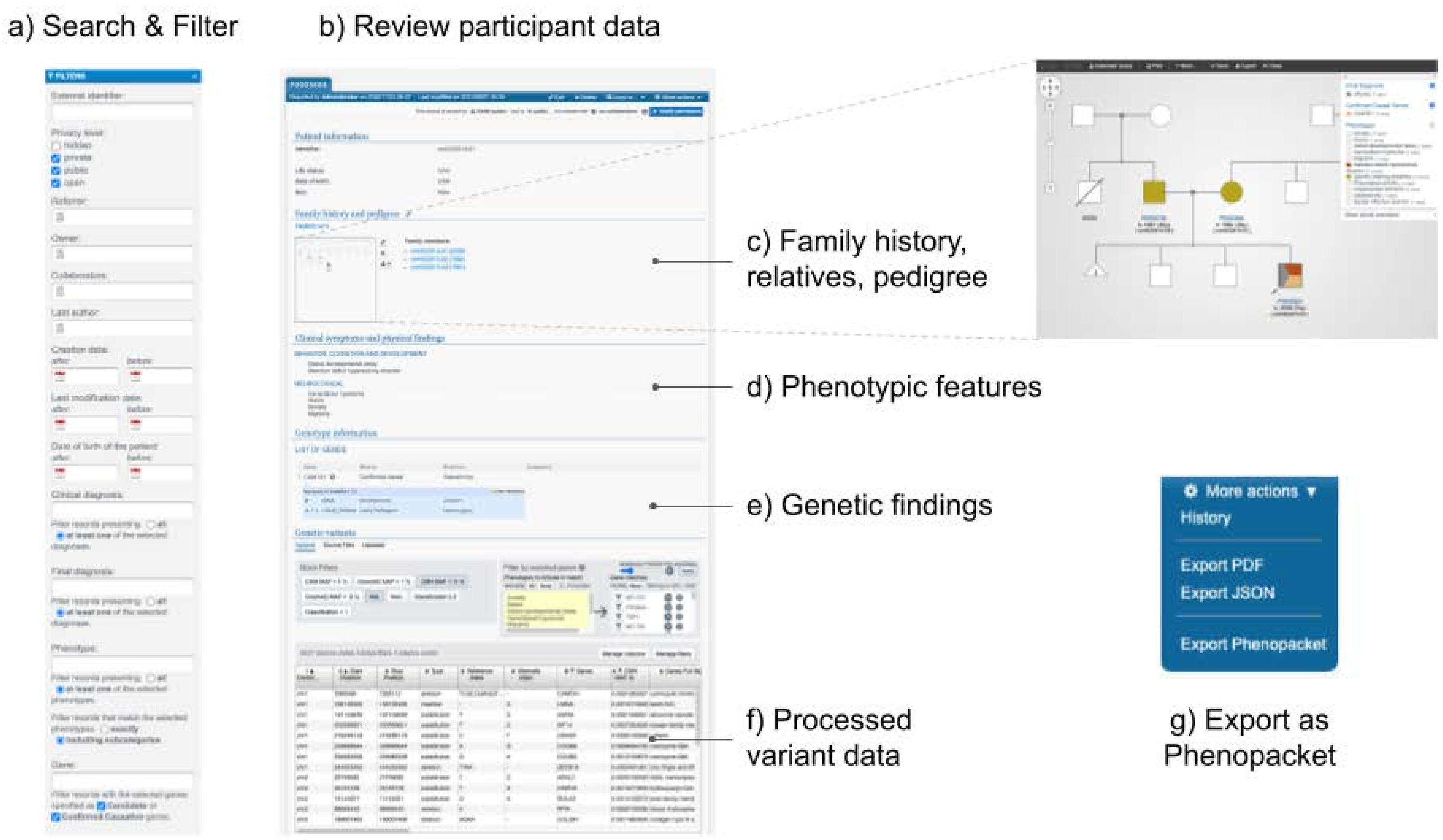
