## Supplemental Methods for "IGenomic answers for children: Dynamic analyses of >1000 pediatric rare disease genomes"

Ana S.A. Cohen et al.

### SUPPLEMENTARY MATERIALS and METHODS

Cohort

Over the initial seven years, "Genomic Answers for Kids" (GA4K) aims to collect genomic data and health information from 30,000 children and their families, ultimately creating a sharable database of nearly 100,000 genomes. Primary recruitment takes place at Children's Mercy Kansas City (CMKC) covering a population base of 5.2M, where the majority of pediatric cases requiring advanced genetic services are seen – 4,000 NGS tests are ordered annually, representing approximately 1500-2000 rare disease families with diverse phenotypes. The clinical catchment extends to seven Midwest states and additional families are referred to our research program from several institutions [IRB 11120514]. Enrollment criteria are intentionally broad and limited only to a suspected underlying genetic diagnosis. Patients with clinically confirmed diagnoses have also been included.

Short-read exome and genome sequencing (ES/srGS)

Exome libraries were prepared according to manufacturer's standard protocols for Illumina TruSeq library preparation (Illumina, San Diego, CA) and IDT XGen Exome Enrichment (IDT, Coralville, IA) following manufacturers' protocols as previously described.[1] Briefly, 250 ng of high quality gDNA is sheared by Covaris sonication (Covaris, Woburn, MA) to an average size of 450 base pairs. After confirmation of the DNA size, the DNA fragments undergo end-repair, a-tailing, adapter ligation and associated AMPure bead (Beckman Coulter, Brea, CA) cleanups according to Illumina's protocol on a Hamilton NGS Star liquid handler (Hamilton Company, Reno, NV). IDT's unique dual indexes with unique molecular identifiers are used. After adapter ligation, 10cycles of standard PCR is performed with Kapa HiFi master mix (Roche, Basel, Switzerland) and primers specific to the ligated adapters, followed by a bead cleanup. The resulting libraries undergo quality assessment for appropriate concentration and final library size. Six libraries are pooled together at 1000ng each to move into IDT's standard exome enrichment. Pooled libraries are blocked to prevent non-specific binding and lyophilized with a vacuum concentrator. Pools are reconstituted with IDT's enrichment buffer, standard exome baits, CNV baits and custom mitochondrial baits. This exome enrichment reaction undergoes a four-hour hybridization according to IDT's standard protocol. The hybridized pool is bound to and incubated with streptavidin beads and then undergoes a series of wash steps with IDT's enrichment wash buffers on a Perkin Elmer Sciclone liquid handler (Perkin Elmer, Watham, MA). An additional 10cycles of standard PCR is performed with Kapa HiFi master mix and primers specific to the adapters, followed by a final bead cleanup. The resulting enriched pools undergo quality assessment for appropriate concentration and final library size, as well as a TaqMan qPCR assay (ThermoFisher, Waltham, MA) to ensure successful enrichment of target regions before standard Illumina Free-Adapter Blocking is performed. Cleaned, adapter-blocked pools are loaded on a NovaSeq6000 with a run configuration of 151x8x8x151. Targeted coverage was a minimum of 80X.Targeted coverage was a minimum of 80X.

PCR-Free whole genome libraries were prepared according to manufacturer's standard protocols for Illumina TruSeq library preparation. Briefly, 1 ug of high quality gDNA is sheared by Covaris sonication an average size of 450 base pairs. After confirmation of the sizes, the DNA fragments undergo end-repair, a-tailing, adapter ligation and associated AMPure bead cleanups according to Illumina's protocol on a Hamilton NGS Star liquid handler. IDT's unique dual indexes with unique molecular identifiers are used. The resulting libraries undergo quality assessment for appropriate concentration and final library size before standard Illumina Free-Adapter Blocking is performed. Cleaned, adapter-blocked libraries are loaded on a NovaSeq6000 with a run configuration of 151x8x8x151. Targeted coverage was a minimum of 30X.

<u>Variant ranking tools</u>

Exomiser version v12.1 (data version 2102) and AMELIE version v3.1.0 were applied for variant prioritization.[2-4] The latter only utilized the intersection of variant calls (categories 1-3) per exome or genome and the gene list produced from the HPO terms for the individual. Category (Cat) 1 variants are those present in HGMD and/or ClinVar; Cat 2 variants are those predicted to cause loss-of-function (nonsense, frameshift, canonical splicing); Cat 3 includes primarily missense variants, but also synonymous variants close to splice junctions. The rationale for not employing the full model (encompassing both HPO and variant modules for AMELIE) was, in part, driven by the desire to not have different emphasis on candidate genes and less weight on variant characteristics. Importantly, the input for Exomiser were unfiltered vcfs, whereas for AMELIE the annotations were based on our internal variant warehouse[1], allowing for putatively different annotations for some variants. Of note for AMELIE, we generated scores only for genes that had at least a single category 1-3 variant at lower than 0.005 population frequency (based on both gnomAD and CMKC internal variant warehouse), meaning that common pathogenic variants for recessive disorders were filtered out (see Supplementary Table S11).

Combined analysis was compiled in "ranked" tsv files (these represent the top 50 overall ranked variants for each case), and high ranks were manually reviewed. This list only includes SNVs and small indels. Expert assessment prioritized X-linked hemizygous variants in males and variants absent in gnomAD for disorders of suspected autosomal dominant inheritance. Additionally, when parents were available and unaffected, *de novo* variants were flagged. For disorders of suspected autosomal recessive inheritance, homozygous variants and two distinct variants in the same gene were prioritized; when parents were available, variants *in cis* were excluded as top candidates.

<u>MGI sequencing (srGS)</u>

Whole genome sequencing of the samples was performed on an MGI DNBSEQ-G400. WGS libraries were constructed using the MGIEasy Universal DNA Library Prep Set according to manufacturer's standard protocols (MGI, Shenzhen, Guangdong, China). A total of 1000 ng of genomic DNA in a 50 ul volume, in micro TUBE-50 AFA Fiber Plate (Covaris #520168) was fragmented using a Covaris ultrasonicator to achieve a length distribution of 100–700 bp with a peak at 400 bp, followed by size selection using AMPure XP (Beckman). The fragmentation product was transferred to a separate tube for end-repair and A-tailing. An equimolar mixed set of MGIEasy PF Barcode Adapters was used for ligation. These ligated DNA fragments were denatured, followed by circularization of the single-stranded DNA, and cleaned up with exonuclease and AMPure XP. After the cleanup of the Exo digestion product, its concentration was measured using a Qubit and normalized to a final concentration of 75 fmol.

Formation of the DNB nanoballs from the circularized ssDNA was carried out according to manufacturer's instructions. The DNB concentration was measured using Qubit with the use of the ssDNA kit. The typical range of nanoball concentrations suitable for loading is 8–40 ng/ µL, however we used a concentration range of 8-20 ng/µL. Nanoball loading onto the flowcell was assisted by a MGI-DL-200RS DNB auto-loader, followed by transfer of the flowcell to the DNBSEQ-G400 for paired-end 150bp sequencing.

PacBio HiFi sequencing (HiFi-GS) and analysis

A total of 8ug of DNA was sheared to a target size of 14 kb using the Diagenode Megaruptor3 (Diagenode, Liege, Belgium) with the following settings: Speed 36, vol 300uL, conc 33ng/uL. A single 1x bead cleanup is completed after shearing using ProNex beads (Promega, Madison, WI), with a final elution volume of 60uL. Library preparation was automated using a Sciclone GX instrument (Perkin Elmer). SMRTbell libraries were prepared with the SMRTbell Express Template Prep Kit 2.0 (100-938-900, Pacific Biosciences, Menlo Park, CA) following the manufacturer's protocol (101-693-800) with modifications. Ligation is completed overnight (20C hold overnight, 65C 10 minutes with a 4C hold). Nuclease v1 is used, with an incubation of 37C for 1 hour. Fragments longer than 10 kb were selected using the Sage Science PippinHT (Sage Science, Beverly, MA). Size-selected libraries were sequenced on the Sequel IIe Systems using Sequel II Binding Kit 2.0 (101-842-900) or 2.2 (102-089-000) and Sequel II Sequencing Kit 2.0 (101-820-200) with 30 hr movies. 175 samples were sequenced to a target of >25X coverage; 297 samples were sequenced on 1 SMRT Cell (average: 10X coverage).

Read mapping, variant calling, and genome assembly were performed using a Snakemake workflow (https://github.com/PacificBiosciences/pb-human-wgs-workflow-snakemake). HiFi reads were mapped to GRCh38 (GCA_000001405.15) with pbmm2 v1.4.0 (https://github.com/PacificBiosciences/pbmm2). Structural variants were called with pbsv 2.4.0 (https://github.com/PacificBiosciences/pbsv) with "--min-gap-comp-id-perc 97.0 --tandem-repeats human_GRCh38_no_alt_analysis_set.trf.bed" options to pbsv discover and "--ccs -m 20 -A 3 -O 3" options to pbsv call. Small variants were called with DeepVariant 1.0 following DeepVariant best practices for PacBio reads (https://github.com/google/deepvariant/blob/r1.2/docs/deepvariant-pacbio-model-case-study.md). [5] *De novo* assembly was performed with hifiasm v0.9-r289 using default parameters.[6]

Structural variant call sets were compared using svpack match (https://github.com/PacificBiosciences/svpack), which considers two SV calls to match when the variants are of the same type (considering INS and DUP to be the same), nearby (start position difference ≤ 100 bp), and similar size (size difference ≤ 100 bp). To systematically evaluate expansions at known pathogenic tandem repeat loci (https://github.com/mcfrith/tandem-genotypes/blob/master/hg38-disease-tr.txt), tandem-genotypes was used to count the length of tandem repeats in HiFi reads for each sample.[7] As long [GA]-rich repeats have been noted to have lower coverage in HiFi reads, a complementary system was setup to identify haplotypes with coverage dropouts at the known pathogenic tandem repeat loci.[8] At each locus, the number of reads that span the repeat region were counted per haplotype (based on a Whatshap-haplotagged BAM from phased SNVs).[9] A coverage dropout was identified as a locus with fewer than 2 spanning reads in a haplotype.

Joint calling of structural and small variants was also completed for HiFi-GS. A multi-sample structural variant callset was produced by merging single-sample pbsv callsets with JASMINE v1.1.4 using jasmine --output-genotypes.[10] A multi-sample small variant callset was produced by running GLnexus v1.2.7 on all single-sample DeepVariant gVCF files using glnexus_cli --config DeepVariant_unfiltered and converting the resulting BCF to VCF with bcftools view v1.10.[11]

<u>10X-linked srGS</u>

10x Linked Read Genome libraries were prepared according to manufacturer's standard protocol (10x Genomics, Pleasanton, CA). Briefly, 5ng of HMW DNA is combined with included reagents and loaded onto a 10x Chromium Genome Chip. The standard Chromium protocol is performed, generating GEMs (Gel Bead-In Emulsions) for each sample. These GEMs undergo an isothermal incubation and a Silane DynaBead (Invitrogen, Waltham, MA) cleanup, followed by a SPRI-Select bead cleanup. Resulting barcoded DNA is quantified and sized with an Agilent TapeStation (Agilent, Santa Clara, CA). Samples of appropriate size and concentration are taken into standard NGS library preparation – End Repair and A-tailing followed by Adapter Ligation. A SPRI-Select bead cleanup is performed before the library proceeds into an Index PCR, with 10 cycles of PCR. A final double-sided SPRI-Select bead cleanup is performed on the finished library. The completed libraries undergo quality assessment for appropriate concentration and final library size before standard Illumina Free-Adapter Blocking is performed. Cleaned, adapter-blocked pools are loaded on a NovaSeq6000 with a run configuration of 151x8x8x151. Targeted coverage was a minimum of 40X. The data generated showed lower quality than PCR-free (Illumina or MGI) srGS when adjusted for sequencing depth and compared against genotyping array QC. Lower replicability of SV calls limited the use of SV data generated from 10X GS and this analysis was omitted.
